# Development and validation of a risk prediction algorithm to estimate all-cause mortality among community-dwelling Canadians – the Mortality Population Risk Tool (MPoRT)

**DOI:** 10.64898/2026.06.12.26355459

**Authors:** Douglas G. Manuel, Anan Bader Eddeen, Philippe Fines, Carol Bennett, Stacey Fisher, Mahsa Jessri, Richard Perez, Meltem Tuna, Claudia Sanmartin, Yulric Sequeria, Juan Li, Laura C. Rosella, Deirdre Hennessy

## Abstract

**Background:** The risk of all-cause mortality can inform decision-making for chronic disease prevention. We developed a predictive algorithm to estimate the 5-year risk of death among community-dwelling adults.

**Methods:** We derived and validated the Mortality Population Risk Tool (MPoRT) using data from population health surveys in Canada (the Canadian Community Health Survey) and the United States (the National Health Interview Survey), survey years 2001 to 2011, linked to vital statistics. The outcome was death within five years of the survey response. The algorithm was developed using data from Ontario respondents using a Cox proportional hazards model, then modified and re-estimated to allow cross-national assessment in Canada and the United States. Twenty-three prespecified predictors were assessed: seven sociodemographic, six behavioural, and ten general health and chronic disease.

**Results:** 527,369 respondents aged 20–105 years were included in the Canadian and United States development and validation cohorts, with 43,758 deaths during 3.68 million person-years follow-up. The final sex-specific MPoRT algorithms each contained 21 variables, showing strong discrimination (C-statistic: females 0.874 [0.871–0.877]; males 0.867 [0.865–0.871]) and good calibration overall and in 246 of 247 subgroups. Discrimination was modestly attenuated (0.01 decrease in C-statistic) in cross-national validation between Canada and the United States, with good calibration across all 71 subgroups.

**Interpretation:** MPoRT accurately discriminated all-cause mortality using only self-reported data, enabling broad application without clinical measures. While validation outside North America is needed to confirm broader applicability, MPoRT is designed for straightforward recalibration using routinely available national mortality data. This supports targeted chronic disease prevention strategies at both the population and individual levels, though the limitations inherent to self-reported predictors should be considered when interpreting predictions.

## Background

Reducing premature mortality and increasing longevity are cornerstones of public health and clinical care.[1–3] Rather than all-cause mortality and overall longevity, the current focus of public health prevention and clinical care decision-making is disease-specific risk assessment, despite many chronic diseases sharing common sociodemographic and preventable risk factors.[4] An all-cause mortality risk assessment could complement disease-specific assessment, especially when modifiable risks or interventions involve multiple chronic conditions and health pathways.[5]

The Global Burden of Disease (GBD) study and clinical risk prediction algorithms exemplify current decision-making practices in population and clinical settings. The GBD study estimates years of life lost for a comprehensive list of over 350 diseases and injuries.[4] It reports the summary burden of preventable risks by estimating and summing risk factor–attributable burdens for individual diseases.[6] In clinical practice, disease-specific risk prediction algorithms are commonly used to guide decision-making. For example, a patient’s baseline risk of developing cardiovascular disease can be estimated using multivariable algorithms like the Framingham Risk Score or the QRISK algorithm.[7,8] These risk estimates inform decisions about the relative, absolute, or lifetime benefits of interventions such as lifestyle modifications or preventive medications.[9] However, these benefits are typically estimated for individual diseases, with only general discussions about the combined benefits of interventions that span multiple conditions, such as improving diet, increasing physical activity, or addressing mental health.[10–12] There is a lack of comprehensive models that empirically assess all-cause mortality risk in clinical settings, which could provide a more holistic approach to patient care.[5,10,13–15]

We hypothesize that all-cause mortality risk can be accurately predicted in the general population using common risk factors collected through population surveys. Assessing all-cause mortality should start with risks known to contribute to mortality across multiple chronic diseases, such as sociodemographic characteristics (e.g., age, sex, socioeconomic status) and health behaviours (e.g., smoking, physical inactivity, poor diet).[6] Health behaviours are associated with more than 50% of all-cause mortality and are important predictors for many chronic diseases.[16–18]

This study aimed to develop and validate a 5-year all-cause mortality risk prediction algorithm using self-reported risk factors commonly collected in population health surveys. The algorithm’s intended purpose is to support population and clinical decision-making by estimating all-cause mortality risk, which supports the estimation of health benefits using absolute or lifetime benefit measures.[19,20] We sought to specify an algorithm that: (1) accurately predicts mortality in general populations; (2) is well-calibrated (unbiased) across a broad range of policy-relevant, clinical, and equity-relevant subgroups; (3) provides both population-level and patient-oriented risk assessments; and (4) can be recalibrated using routinely published national actuarial life tables.

## Methods

The predictive algorithm—the Mortality Population Risk Tool (MPoRT)—was developed using routinely-collected population health survey data linked at the individual level to all-cause mortality. The initial algorithm development and temporal validation were based on a similar study[21] and data (the Ontario sample of the Canadian Community Health Survey (CCHS)). The national sample of CCHS and the United States (US) National Health Interview Survey (NHIS) became available after the development and temporal validation of MPoRT. These data provided the opportunity for further development and external validation of the algorithm in two nationally representative samples. assessment of recalibration using reference life tables created by national statistical agencies (Statistics Canada and the US National Center for Health Statistics).[22]

### Data sources

#### Development cohort

The Ontario samples of the CCHSs conducted in 2001, 2003, 2005, and 2007-2008 were used for the development cohort. Respondents were followed from the survey administration date until the earliest of death, loss to follow-up (defined as loss of health care eligibility), or end of study (April 30, 2016).

The CCHS is a cross-sectional survey conducted by Statistics Canada that collects data related to health determinants, health status and healthcare use among household dwellers. The survey uses multistage sampling to select households in each health region randomly. Details of the survey methodology have previously been published.[23] A survey weight, which reflects the number of individuals represented in the target population, is assigned to each respondent. The CCHS target population includes individuals aged 12 and older living in Canada’s ten provinces and three territories. Individuals living in First Nation Reserves, institutionalized residents, full-time Canadian Forces members, and residents of certain remote areas are excluded from the sampling frame.

#### Validation cohorts

The Ontario samples of the 2009-2010 and 2011-2012 CCHS cycles were used as a temporal validation cohort. External validation was performed using the national (excluding Ontario) 2003, 2005 and 2007-2008 CCHS cycles and the 2000 and 2005 cycles of the NHIS, linked to individual-level mortality with follow-up to December 31, 2011. The NHIS had a similar design, target populations and content to the CCHS.[24,25]

### Participants

Survey respondents in the development and validation cohorts were included if they were aged 20 years or older. For individuals with multiple interviews, only the earliest interview was included. In the development cohort, respondents were excluded if they were not eligible for Ontario’s universal health insurance program (required to link survey responses to vital statistics in Ontario) or did not agree to share their survey responses.

### Outcome

The outcome, all-cause mortality, was ascertained in the development and temporal validation cohorts using Vital Statistics and the Registered Persons Database (Ontario). In the external validation data, Statistics Canada linked the national CCHS survey respondents to the Canadian Vital Statistics Database[26]. The National Center for Health Statistics linked the NHIS respondents to the National Death Index.[17]

### Predictors

Predictors were ascertained using self-reported responses from the CCHS and NHIS surveys. All predictors and interaction terms were pre-specified before algorithm development and maintained in their original form (i.e., categorical or continuous).

Separate sex-specific MPoRT algorithms were developed. The development cohort included 23 predictor variables: seven socio-demographic (age, sex, ethnicity, immigrant status, fraction of lifetime in Canada, education, and neighbourhood deprivation), six behavioural (smoking status, pack years of smoking, alcohol consumption, former/non-drinker status, simplified diet score, and leisure time physical activity), and ten general health and chronic condition variables (diabetes, high blood pressure, chronic respiratory conditions, mood disorders, active cancer, dementia, heart disease, stroke, epilepsy, and body mass index). The model included interactions between age and all other predictors, except immigrant status, fraction of lifetime in Canada (for the development algorithm) and ethnicity. **Table 1** describes how these variables were defined and modelled.

**Table 1.** Predictor variables for the Mortality Population Risk Tool (MPoRT)

| Variable | Scale | Initial variable specification | Full model<br>Model degrees of freedom: 64 (female); 63 (male) | Reduced model<br>Model degrees of freedom: 61 (female); 60 (male) |
| --- | --- | --- | --- | --- |
| **Demographic** |  |  |  |  |
| Age* | Continuous | **5 knot spline**<br>Valid range: 20 to 102 (female & male) | Unchanged | Unchanged |
| Sex | Dichotomous | Stratified: female, male | n/a | n/a |
| **Health behaviours** |  |  |  |  |
| Pack years of smoking | Continuous | **3 knot spline**<br>Valid range: 0 to 78 (female), 0 to 112.5 (male) | Unchanged | Unchanged |
| Smoking status | Categorical | **4 categories:**<br>• Non-smoker<br>• Current smoker<br>• Former ≤5 years<br>• Former >5 years | Unchanged | Unchanged |
| Alcohol<br>(number of drinks past week) | Continuous | **4 knot spline** (female)<br>**3 knot spline** (male)<br>Valid range: 0-24 (female), 0-50 (male) | Unchanged | Unchanged |
| Former/non-drinker | Dichotomous | Yes/No | Unchanged | Unchanged |
| Simplified diet score<sup>†</sup> | Continuous | **3 knot spline**<br>Valid range: -18.9 to 20.7 (female), -16.8 to 18.4 (male) | Unchanged | Unchanged |
| Leisure physical activity<br>(average daily Metabolic Equivalent of Task [MET]) | Continuous | **3 knot spline**<br>Valid range: 0 to 12.4 (female), 0 to 16 (male) | Unchanged | Unchanged |
| **Socio-demographic** |  |  |  |  |
| Ethnicity | Categorical | **6 categories:**<br>• White<br>• Black<br>• Chinese<br>• South Asian; Arab; West Asian<br>• Japanese; Korean; Southeast Asian; Filipino<br>• Other; Indigenous; multiple origin; unknown; Latin American | Unchanged | Unchanged |
| Immigrant | Dichotomous | Yes, No | Unchanged | Excluded |
| Fraction of lifetime in Canada | Continuous | **3 knot spline**<br>Valid range: 0 to 1 | Unchanged | Excluded (male), unchanged (female) |
| Education | Categorical | **4 categories:**<br>• Less than secondary<br>• Secondary school graduation<br>• Some post-secondary<br>• Post-secondary graduation | Unchanged | Unchanged |

<sup>†</sup> Pampalon R, Hamel D, Gamache P, Raymond G. A deprivation index for health planning in Canada. Chronic Dis Can. 2009 Jan 1;29(4):178-91.

| Variable | Scale | Initial variable specification | Full model | Reduced model |
| --- | --- | --- | --- | --- |
| Neighbourhood social and material deprivation (Pampalon's deprivation index<sup>1</sup>) | Ordinal | **3 categories:**<br>Low (1<sup>st</sup> or 2<sup>nd</sup> quintile);<br>High (4<sup>th</sup> or 5<sup>th</sup> quintile);<br>Moderate (all others) | Unchanged | Unchanged |
| **Chronic conditions** |  |  |  |  |
| Diabetes | Dichotomous | Yes, No | Unchanged | Unchanged |
| High blood pressure | Dichotomous | Yes, No | Unchanged | Unchanged |
| COPD, Emphysema, Chronic Bronchitis | Dichotomous | Yes, No | Unchanged | Unchanged |
| Mood disorders | Dichotomous | Yes, No | Unchanged | Unchanged |
| Cancer (active)<sup>‡</sup> | Dichotomous | Yes, No | Unchanged | Unchanged |
| Dementia | Dichotomous | Yes, No | Unchanged | Unchanged |
| Heart disease | Dichotomous | Yes, No | Unchanged | Unchanged |
| Stroke | Dichotomous | Yes, No | Unchanged | Unchanged |
| Epilepsy | Dichotomous | Yes, No | Unchanged | Excluded (female),<br>unchanged (male) |
| Body mass index | Continuous | **3 knot spline**<br>Valid range: 8.9 to 47.2<br>(female), 8.6 to 43.7<br>(male) | Unchanged | Unchanged |

\*Age interaction included for all variables except immigrant, fraction of time in Canada and ethnicity
<sup>†</sup>Simplified diet score: Prior to summation, all diet variables were rescaled (mean = 0, standard deviation = 1). The simplified diet score was calculated by summing the daily reported intake frequency of all diet variables (fruit, vegetables (including carrots, salad and potatoes), and fruit juice) and subtracting any potato and fruit juice intake frequency greater than one (overconsumption penalty)
<sup>‡</sup>Cancer diagnosed within the last 5 years

The predictors were pre-specified and harmonized across surveys for the national validation data. For example, the NHIS did not ask questions to calculate pack-years of smoking. Therefore, we simplified the smoking exposure to a consistent exposure of ‘heavy’ (greater or equal to one pack (20 cigarettes) of smoking per day) and ‘light’ smoker status (less than one pack per day). We identified additional sociodemographic variables available in the CCHS (Canada) and NHIS (United States), including marital status, home ownership and income ratio. **Additional file 1** compares the predictors in the national external validation models to the Ontario reduced model.

### Statistical methods and analysis

The pre-specified protocol for the development and validation of MPoRT followed the methods of a previously published cardiovascular risk algorithm—the Cardiovascular Population Risk Tool (CVDPoRT) (clinicaltrial.org NCT02267447)—and an earlier all-cause and premature mortality algorithms.[27,28,17]

The protocol followed the Transparent Reporting of a Multivariable Prediction Model for Individual Prognosis or Diagnosis (TRIPOD) reporting guidelines[29] and adhered to additions in the updated TRIPOD-AI version.[30] We followed the approach of Harrell[31] and Steyerberg[32], including developing the analysis plan before any model fitting or descriptive analyses involving exposure-outcome associations, fully pre-specifying the predictor variables, using flexible functions for continuous predictors, and preserving statistical properties by avoiding data-driven variable selection procedures.

Our approach for development of MPoRT in the Ontario population adhered to the CVDPoRT protocol [27] with the following exceptions: study data included additional CCHS cycles (2009-1010 and 2011-2012), follow-up time was to April 30, 2016 (versus December 31, 2012); pregnant respondents and those with pre-existing cardiovascular disease were included; the outcome was all-cause mortality (as opposed to cardiovascular disease); additional chronic conditions were added (chronic respiratory conditions, mood disorders, active cancer, dementia, heart disease, stroke, and epilepsy); diet predictors were modified to generate a simplified diet score; and, two general health predictors were not assessed (self-perceived stress and sense of belonging to the local community) due to concerns about generalizability in other settings and because of low predictive ability in CVDPoRT.

As discussed, additional sociodemographic predictors were added for the nationally derived algorithms in Canada and the US. These predictors were included after the derivation of the Ontario algorithm but before analyses of the predictor-outcome relationship in Canada-wide and United States data. We re-derived the Ontario reduced model separately in each cohort, using the simplified, harmonized predictors for the national Canadian and United States cohorts. This means that we generated two additional algorithms: one using Canadian data and another using United States data. Following this, the performance of the Canadian model was validated in the United States cohort and vice versa. Finally, we combined the national Canadian and United States cohorts to create and validate a combined two-country algorithm.

Data cleaning, model specification and model estimation are presented in **Additional file 2**. Briefly, we used multiple imputations to impute missing values on predictor variables.[31] Models were estimated using Cox proportional hazards after assessing the proportionality assumption. A preliminary main effects model was fit that included a pre-specified initial degree of freedom allocation for each predictor.[31,33] We used the step-down procedure described by Ambler et al. to identify a parsimonious model for applications where parsimony may be more important than accuracy.[34] This procedure involves sequentially removing variables that minimally impact model fit to a desired degree of accuracy based on their contribution to the model’s R^2^. [34]

Performance in the derivation and validation cohorts was assessed using discrimination measures (ability to differentiate risk between individuals) and calibration (agreement between predicted and observed risk). Discrimination was assessed using the C-statistic and the ratio of the 95 to 5 risk percentiles. Calibration was assessed using calibration plots, overall observed (death) versus predicted (risk), and observed versus predicted by specific subgroups. Subgroups were examined using predefined criteria that were clinically or policy relevant (<20% difference between observed and predicted estimates for categories with a prevalence higher than 5%).[35,36] The scaled Brier Score and Nagelkerke R^2^ were calculated for overall performance measures.[19,26]

### Survey re-weighting to national mortality risk

Despite using population-based surveys, we expected differences in selection, response and linkage biases between the surveys. This could result in different mortality risks estimated from the study data compared to the official national life expectancy estimates. However, the availability of national mortality risk estimates enables recalibration of all-cause mortality risk algorithms to improve their generalizability and accuracy in real-world applications. Updating and recalibrating models using age- and sex-specific groups provides greater specificity than the more common approach of recalibrating a model in-the-whole.[37–39]

In anticipation of this recalibration approach, we generated an additional calibration offset before deriving the Canada and United States models based on the observed relative differences between the age- and sex-specific mortality rates calculated from the national vital statistics mortality rates.

## Results

### Participants

There were 527,369 respondents in the derivation and validation cohorts, 43,758 died over 3.68 million person-years of follow-up. After exclusions, the Ontario derivation cohort comprised 116,030 unique respondents with 1.13 million person-years of follow-up and 15,315 deaths; the Ontario validation cohort had 56,709 unique respondents with 237,000 person-years of follow-up and 2,875 deaths. The national CCHS external validation cohort included 296,407 respondents with 1.81 million person-years of follow-up and 19,227 deaths; the NHIS external validation cohort included 58,223 respondents with 0.5 million person-years of follow-up and 6,341 deaths. (**Additional file 3** and **Additional file 4**) **Additional file 5** (Ontario derivation and validation cohorts) and **Additional file 6** (external validation cohorts) present characteristics of the study populations. Several predictors weren’t asked of respondents in all survey years: chronic respiratory conditions, mood disorders, dementia, epilepsy, and former drinker status. Apart from these, less than 5% of the predictors had missing data.

### Model development and specification

Separate sex-specific MPoRT algorithms were developed. For the female model, the development cohort had 63,470 participants and 7,969 deaths. For the male model, there were 52,560 participants and 7,346 deaths. The predictors and their initial and final degrees of freedom are shown in **Table 1**. In adherence with our protocol, we did not examine unadjusted associations between candidate predictors and mortality outcomes. **Table 2** presents adjusted hazard ratios for the MPoRT final reduced models. The complete model specifications, estimated coefficients, and model formula are in **Additional file 1, Additional file 7, and Additional file 8.**

**Table 2.** Adjusted hazard ratios for five-year risk of death from the Mortality Population Risk Tool (MPoRT) Ontario reduced models.

| Predictor | Adjusted hazard ratio (95% confidence interval) |  |
| --- | --- | --- |
|  | Female model | Male model |
| **Age<sup>†</sup>** |  |  |
| 80 | 12.57 (6.74–23.47) | 23.51 (12.83–43.10) |
| 20 | 0.07 (0.02–0.19) | 0.15 (0.06–0.40) |
| Mean age | 1.00 | 1.00 |
| **Smoking<sup>‡</sup>** |  |  |
| Current | 2.03 (1.80–2.29) | 2.24 (1.96–2.55) |
| Former smoker ≤5 years | 1.75 (1.50–2.04) | 1.59 (1.35–1.87) |
| Former smoker >5 years | 1.29 (1.14–1.46) | 1.23 (1.07–1.40) |
| Non-smoker | 1.00 | 1.00 |
| **Alcohol** |  |  |
| Drinks previous week |  |  |
| 21 | 1.43 (1.16–1.77) | 1.02 (0.94–1.10) |
| 5 | 1.04 (0.97–1.12) | 0.93 (0.91–0.96) |
| 2 | 1.00 | 1.00 |
| Former/non-drinker | 1.20 (1.08–1.33) | 1.22 (1.10–1.37) |
| Current drinker | 1.00 | 1.00 |
| **Simplified diet score (percentile)<sup>§</sup>** |  |  |
| 10<sup>th</sup> | 1.09 (0.99–1.21) | 1.12 (1.01–1.24) |
| 30<sup>th</sup> | 1.07 (0.99–1.16) | 1.08 (1.00–1.17) |
| 50<sup>th</sup> | 1.05 (0.99–1.12) | 1.05 (0.99–1.12) |
| 70<sup>th</sup> | 1.03 (0.99–1.08) | 1.03 (0.99–1.08) |
| 90<sup>th</sup> | 1.00 | 1.00 |
| **Physical activity (average kcal/kg/day)** |  |  |
| 0 | 1.70 (1.55–1.86) | 1.45 (1.34–1.58) |
| 1 | 1.31 (1.24–1.38) | 1.23 (1.17–1.29) |
| 2 | 1.08 (1.06–1.11) | 1.07 (1.05–1.10) |
| 3 | 1.00 | 1.00 |
| **Ethnicity** |  |  |
| Black | 1.01 (0.77–1.34) | 0.86 (0.64–1.15) |
| Chinese | 0.58 (0.41–0.81) | 0.61 (0.47–0.79) |
| South Asian; Arab; West Asian | 0.69 (0.51–0.93) | 0.48 (0.37–0.61) |
| Japanese; Korean; Southeast Asian; Filipino | 0.70 (0.51–0.97) | 0.72 (0.53–0.99) |
| Other | 0.95 (0.82–1.10) | 0.94 (0.82–1.08) |
| White | 1.00 | 1.00 |
| **Education** |  |  |
| Less than secondary school graduation | 1.15 (1.04–1.28) | 1.18 (1.07–1.30) |
| Secondary school graduation | 1.05 (0.94–1.16) | 1.04 (0.93–1.15) |
| Some post-secondary education | 1.01 (0.86–1.19) | 1.03 (0.88–1.21) |
| Post-secondary graduation | 1.00 | 1.00 |
| **Neighbourhood deprivation** |  |  |
| High | 1.23 (1.08–1.41) | 1.36 (1.19–1.55) |
| Moderate | 1.07 (0.95–1.20) | 1.09 (0.98–1.22) |
| Low | 1.00 | 1.00 |
| **Diabetes status** |  |  |
| Yes diabetes | 1.90 (1.67–2.17) | 1.92 (1.70–2.16) |
| No diabetes | 1.00 | 1.00 |
| **Hypertension status** |  |  |
| Yes hypertension | 1.15 (1.04–1.27) | 1.17 (1.06–1.29) |
| No hypertension | 1.00 | 1.00 |
| **Respiratory status** |  |  |
| Yes respiratory condition | 1.64 (1.46–1.84) | 1.46 (1.27–1.67) |
| No respiratory condition | 1.00 | 1.00 |
| **Mood disorder status** |  |  |
| Yes mood disorder | 1.14 (1.02–1.28) | 1.42 (1.26–1.61) |
| No mood disorder | 1.00 | 1.00 |
| **Active cancer status** |  |  |
| Yes active cancer | 5.00 (4.39–5.69) | 3.61 (3.08–4.24) |
| No active cancer | 1.00 | 1.00 |
| **Dementia status** |  |  |
| Yes dementia | 3.71 (2.58–5.33) | 2.02 (1.42–2.88) |
| No dementia | 1.00 | 1.00 |
| **Heart disease status** |  |  |
| Yes heart disease | 1.57 (1.38–1.79) | 1.50 (1.32–1.69) |
| No heart disease | 1.00 | 1.00 |
| **Stroke status** |  |  |
| Yes stroke | 1.77 (1.43–2.20) | 1.49 (1.20–1.84) |
| No stroke | 1.00 | 1.00 |
| **Epilepsy status** |  |  |
| Yes epilepsy | n/a | 1.96 (1.48–2.58) |
| No epilepsy | n/a | 1.00 |
| **Body Mass Index (kg/m<sup>2</sup>)** |  |  |
| 40 | 1.21 (1.08–1.35) | 1.39 (1.22–1.59) |
| 30 | 0.95 (0.92–0.99) | 0.96 (0.92–1.00) |
| 25 | 1.00 | 1.00 |

† Predictors with age interaction shown for mean age. Mean age = 52.2 years for females and 50.6 years for males. Age interaction included for all variables except immigrant, fraction of time in Canada and ethnicity. See interactive plot for ages 20 to 80 years.
‡Smoking hazards shown for 10 pack-years.
§Simplified diet score: Prior to summation, all diet variables were rescaled (mean = 0, standard deviation = 1). The simplified diet score was calculated by summing the daily reported intake frequency of all diet variables (fruit, vegetables (including carrots, salad and potatoes), and fruit juice) and subtracting any fruit juice intake and potato intake greater than one (overconsumption penalty)

### Model performance

**Table 3** presents indicators of model performance. The overall C-statistic in the Ontario validation cohort was 0.88 for females (95% CI 0.88–0.89); 0.88 for males (95% CI 0.88–0.89), a slight reduction in discrimination from the development cohorts. The national algorithms had a 0.1 reduction in the C-statistic in the cross-nation validation compared to the original Ontario validation.

**Table 3.** Summary statistics showing goodness of fit for the Mortality Population Risk Tool (MPoRT): development, internal validation, and external validation.

|  | Development | Internal validation |  |  | External validation |  |  |
| --- | --- | --- | --- | --- | --- | --- | --- |
| **Model** | Ontario development | Ontario validation | Ontario combined | Ontario reduced | United States reduced | Canada reduced | Combined United States/Canada |
| **Dataset** | Ontario CCHS 2000–2001, 2003, 2005, and 2007–2008 | Ontario CCHS 2009–2010 and 2011–2012 | Ontario CCHS 2000–2001, 2003, 2005, 2007–2008, 2009–2010 and 2011–2012 | Ontario CCHS 2000–2001, 2003, 2005, 2007–2008, 2009–2010 and 2011–2012 | National CCHS 2003, 2005, 2007–2008 | NHIS 2000 and 2005 | National CCHS 2003, 2005, 2007–2008, NHIS 2000, and 2005 |
| **Female model** |  |  |  |  |  |  |  |
| **Discrimination** |  |  |  |  |  |  |  |
| C–statistic (95% CI) | 0.884<br>(0.880–0.887) | 0.879<br>(0.872–0.880) | 0.883<br>(0.880–0.887) | 0.883<br>(0.880–0.887) | 0.871<br>(0.867–0.873) | 0.867<br>(0.862–0.879) | 0.874<br>(0.871–0.877) |
| Ratio of 95 to 5 risk percentile | 360<br>(0.001–0.260) | 492<br>(0.001–0.264) | 377<br>(0.001–0.263) | 377<br>(0.001–0.263) | 332<br>(0.0009–0.326) | 221<br>(0.002–0.403) | 189<br>(0.002–0.344) |
| **Calibration** |  |  |  |  |  |  |  |
| Observed versus predicted | 0.98% | 3.60% | -0.26% | -0.27% | -2.64% | 3.82% | 1.65% |
| 5-year cumulative incidence (observed) (95% CI) | 0.051<br>(0.049–0.053) | 0.054<br>(0.051–0.057) | 0.052<br>(0.050–0.053) | 0.052<br>(0.050–0.053) | 0.043<br>(0.042–0.044) | 0.051<br>(0.048–0.053) | 0.044<br>(0.043–0.045) |
| 5-year risk (predicted) | 0.050 | 0.052 | 0.052 | 0.052 | 0.045 | 0.049 | 0.044 |
| Calibration slope/intercept | 0.988/0.001 | 1.026/0.0006 | 0.996/0.0004 | 0.995/0.0004 | 0.945/0.001 | 1.058/-0.0009 | 1.002/0.0005 |
| **Overall performance** |  |  |  |  |  |  |  |
| Brier<sub>scaled</sub> Score | 0.201 | 0.201 | 0.206 | 0.206 | 0.148 | 0.192 | 0.163 |
| Nagelkerke R<sup>2</sup> | 0.282 | 0.175 | 0.240 | 0.240 | 0.165 | 0.243 | 0.275 |
| **Male model** |  |  |  |  |  |  |  |
| **Discrimination** |  |  |  |  |  |  |  |
| C–statistic (95% CI) | 0.880<br>(0.876–0.884) | 0.869<br>(0.860–0.877) | 0.878<br>(0.875–0.882) | 0.878<br>(0.875–0.882) | 0.863<br>(0.860–0.867) | 0.855<br>(0.848–0.861) | 0.867<br>(0.865–0.871) |
| Ratio of 95 to 5 risk percentile | 276<br>(0.001–0.322) | 389<br>(0.001–0.333) | 295<br>(0.001–0.327) | 296<br>(0.001–0.327) | 194<br>(0.002–0.405) | 116<br>(0.004–0.459) | 163<br>(0.003–0.416) |
| **Calibration** |  |  |  |  |  |  |  |
| Observed versus predicted | 1.03% | 1.35% | -0.56% | -0.54% | -4.23% | 3.79% | 1.63% |
| 5-year cumulative incidence (observed) (95% CI) | 0.064<br>(0.062–0.067) | 0.069<br>(0.065–0.072) | 0.065<br>(0.064–0.067) | 0.065<br>(0.064–0.067) | 0.056<br>(0.054–0.057) | 0.061<br>(0.057–0.063) | 0.056<br>(0.055–0.0457) |
| 5-year risk (predicted) | 0.064 | 0.068 | 0.066 | 0.066 | 0.058 | 0.058 | 0.055 |
| Calibration slope and intercept | 1.000/0.0003 | 0.989/0.0021 | 0.995/0.0005 | 0.996/0.0004 | 1.054/-0.0056 | 1.012/0.0017 | 1.032/-0.0008 |
| **Overall performance** |  |  |  |  |  |  |  |
| Brier<sub>scaled</sub> Score | 0.240 | 0.240 | 0.245 | 0.245 | 0.183 | 0.211 | 0.193 |
| Nagelkerke R<sup>2</sup> | 0.294 | 0.183 | 0.253 | 0.253 | 0.177 | 0.247 | 0.295 |

Abbreviations: CI = confidence interval, CCHS = Canadian Community Health Survey, NHIS = National Health Interview Survey
Three types of performance tests were examined(1):
**1. Discrimination** is the ability of a prediction model to differentiate between those who do and do not develop the outcome of interest.
**C-statistic** is a rank order statistic for predictions against true outcomes.(2,3) The statistic ranges from 0 to 1: a value of 0.5 indicates the model is no better than random prediction, a value of 1 indicates the model perfectly predicts those who will develop the outcome of interest and those who will not.
**Ratio of 95 to 5 risk percentiles** is a test of discrimination. A higher ratio indicates a more discriminating algorithm. For example, a ratio of 100 indicates that the absolute risk is 100 times higher for a person in the 95<sup>th</sup> percentile than for a person in the 5<sup>th</sup> percentile. The ratio can be used to gauge the potential absolute benefit of treatment for different individuals in the development and validation cohorts. For an intervention with the same relative benefit, a risk ratio of 100 indicates that one person will have 100 times the absolute benefit of the comparative person.
**2. Calibration** reflects agreement between the observed outcomes and predictions. Calibration (or accuracy) describes how well the predicted probability of disease agrees with the observed outcome.

Predicted risk closely approximated observed risk across all validation datasets, with calibration slopes near one and intercepts near zero, indicating good agreement between predicted and observed risks. For females, the calibration slope and intercept were 1.026; 0.0006 (Ontario), 0.945; 0.001 (US), and 1.058; –0.0009 (Canada). For males, values were 0.989; 0.0021 (Ontario), 1.054; –0.0056 (US), and 1.012; 0.0017 (Canada)

(**Figure 1**).

**Figure 1.**
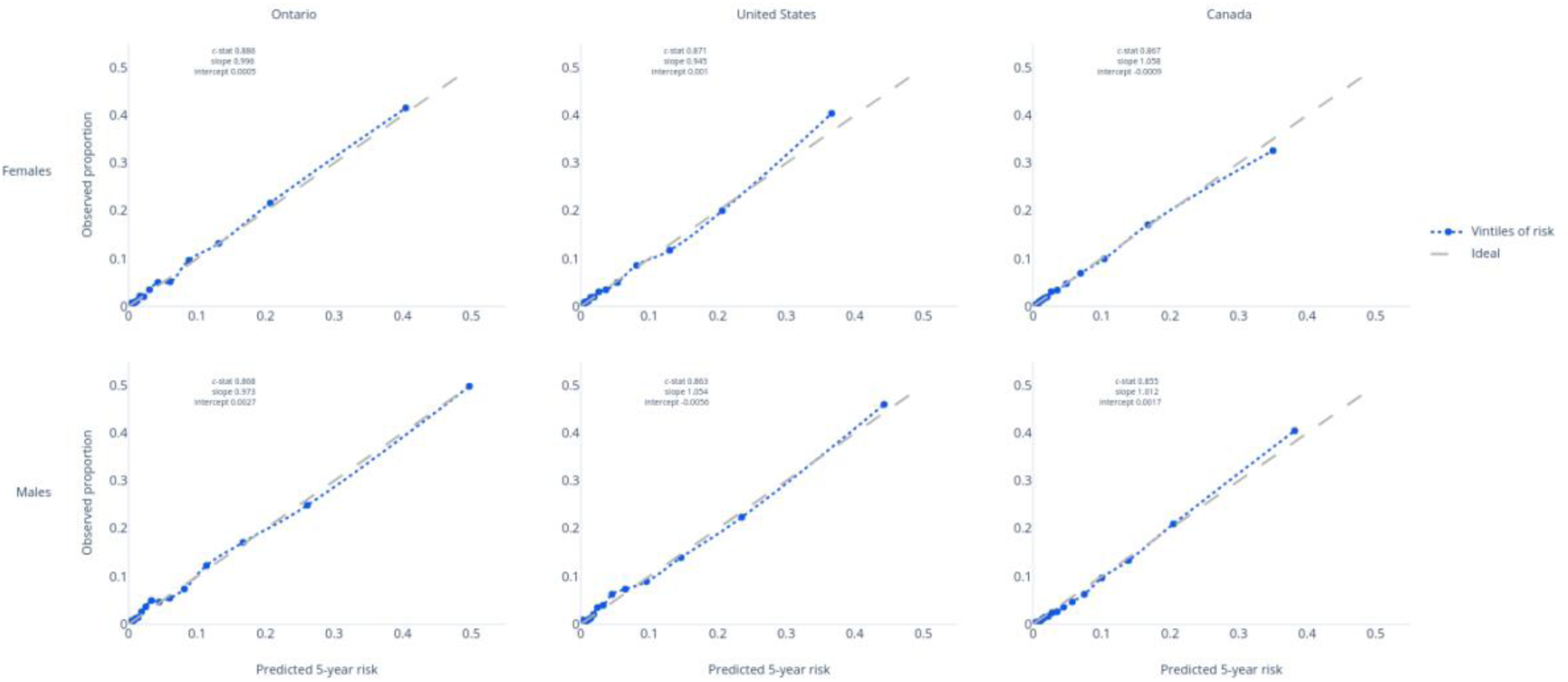
Calibration of the Mortality Population Risk Tool (MPoRT), Ontario, United States and Canada validation data.

Among Ontario females, the algorithm was well-calibrated in 122 of 123 predefined clinically or policy-relevant subgroups, with observed versus predicted risk being greater than the predefined difference of 20% only for those with active cancer (**Additional file 9**). Among Ontario males, the algorithm was well-calibrated in all 124 predefined policy-relevant subgroups (**Additional file 9**). **Additional file 10** and **Additional file 11** present observed versus predicted risk for the national validation cohorts. The algorithm was well-calibrated in all 71 predefined policy-relevant subgroups for both males and females.

The final algorithm will be openly available in Predictive Modelling Mark-up Language (PMML) format on GitHub.

## Discussion

Our study demonstrates that the Mortality Population Risk Tool (MPoRT) can accurately predict all-cause mortality in general populations using self-reported data from large, population-based health surveys. MPoRT offers a practical and accessible approach to all-cause mortality risk assessment, using only self-reported sociodemographic characteristics, health behaviours, and chronic disease status.

### Study strength

The study’s strengths include large development and validation cohorts, external validation using national data in Canada and the United States, and provisions for straightforward recalibration using routinely reported national vital statistics, with or without national health surveys.

We observed that MPoRT maintained high discrimination after modest respecification to accommodate differences in the similar linked population surveys in Canada and the United States. This cross-national validation suggests that MPoRT is robust and potentially generalizable to other populations with similar health survey data.

### Implications for clinical practice and population planning

MPoRT supports decision-making in both clinical and population health settings by providing an overall assessment of an individual’s mortality risk. Unlike disease-specific risk assessments, all-cause mortality estimation captures the cumulative impact of modifiable risk factors, such as health behaviours and sociodemographic characteristics, simultaneously affecting multiple chronic diseases. Estimating risk separately for multiple conditions like cardiovascular disease, diabetes, and cancer is burdensome and rarely performed in practice, making it challenging to fully evaluate the health benefits of interventions targeting risks like smoking, obesity, poor diet, and physical inactivity.

In clinical practice, MPoRT offers a streamlined approach by consolidating multiple risk factors into a single mortality risk estimate using self-reported risk factors. This enables healthcare providers to identify high-risk individuals more efficiently and prioritize interventions with broad impacts across various conditions. For example, a clinician can use MPoRT to quantify the overall mortality benefit of smoking cessation for a patient rather than estimating separate risks for each of over 20 smoking-related diseases.

In population health practice, MPoRT informs public health by projecting future mortality, assessing mortality risk distribution across subgroups, and evaluating preventive strategies and policies.[20] Multivariate risk algorithms like MPoRT provide discriminative estimates of population risk, which Geoffrey Rose identified as crucial for evidence-based prevention program planning.[40]

Furthermore, MPoRT facilitates the assessment of health inequities by incorporating sociodemographic factors into risk estimation. By identifying high-risk groups, public health planners can tailor interventions and target resources toward vulnerable populations, reducing health disparities and improving overall population health.[2, 3]

All-cause mortality risk also provides essential data for calculating additional metrics used in chronic disease decision-making. These include years living with disease, years of life gained, and quality-adjusted life years resulting from prevention or treatment. Estimating these measures requires an accurate assessment of an individual’s life expectancy, which can be more precisely determined using multivariable approaches like MPoRT rather than relying solely on age- and sex-specific averages.

### Caution on interpretation and use

MPoRT is a predictive algorithm designed exclusively to estimate baseline all-cause mortality risk. Its regression coefficients represent statistical associations, not causal relationships. Therefore, MPoRT should not be directly used to quantify specific risk factors’ causal impact or burden. Instead, when assessing the preventable burden of risk factors or potential benefits of interventions, MPoRT-derived baseline risks should be combined with separately obtained, causally informed estimates, such as relative hazards derived from meta-analyses, randomized trials, or expert consensus. This combined approach ensures accurate and interpretable assessments of potential intervention impacts on population health.

### Comparison with existing models

While numerous risk prediction algorithms exist for specific conditions,[41,42] models predicting all-cause mortality in general populations are less common. The Ubble algorithm developed by Ganna and Ingelsson[15] was among the first large-scale studies in the UK to predict all-cause mortality using a data-driven approach with 655 predictors, including self-rated health. Deelen et al.[43] examined the predictive ability of metabolomics for all-cause mortality.

In comparison, MPoRT employs a pre-specified approach using established predictors. When evaluating predictive performance, MPoRT demonstrated higher discrimination than the Ubble and Deelen et al. models. Specifically, MPoRT achieved a C-statistic of 0.88, compared to 0.80 for the Ubble model and 0.79 for the Deelen et al. model. Moreover, MPoRT maintained calibration in external validation, indicating that the predicted probabilities closely matched observed outcomes across different populations. In contrast, the Ubble and Deelen et al. models did not assess calibration within their study populations or external populations, limiting the understanding of their absolute risk predictions in different populations.

### Methods considerations

MPoRT’s development offers several advantages. By pre-specifying predictors based on well-established epidemiological relationships with mortality,[44] the model reduces the potential for overly optimistic predictions that may not be maintained in different settings.

We intentionally excluded self-rated health from MPoRT, despite its strong predictive ability demonstrated in Ganna and Ingelsson.[15] This decision enhanced the model’s cross-cultural applicability, as self-rated health can vary significantly due to cultural and ethnic differences in health perception and reporting.[45] For example, individuals in the United States are more likely to report very good or excellent health than those in Japan, despite Japan’s higher life expectancy.[46,47] Including self-rated health could, therefore, reduce the model’s portability and accuracy in diverse populations.

While we included sociodemographic variables that might also be subject to cultural differences, these factors are generally more objective and can be standardized using internationally recognized classifications. Nonetheless, cultural variability may still influence these predictors and their relationships with mortality.[48]

### Potential for incorporating additional predictors

There is potential to enhance MPoRT by incorporating additional predictors such as genetic and biological markers, environmental exposures, and healthcare access variables. For instance, genomics and metabolites identified in studies like Deelen et al.[43] may capture different mortality pathways not reflected by health behaviours and sociodemographic factors. Including these biomarkers could improve personalized risk assessments, especially for individuals with uncommon risk factors.

However, given MPoRT’s already high discrimination, these additional predictors would need substantial relative contributions or be widely prevalent to improve predictive accuracy meaningfully.[49] Practical considerations such as data availability, cost, and ethical implications must be addressed when considering the inclusion of genetic and metabolomic data.[49–51] Machine learning algorithms could potentially improve predictive performance by capturing complex relationships among predictors.[52] However, we included previously identified interactions and non-linear relationships, and more complex modelling may introduce challenges related to model interpretability and the risk of overfitting, necessitating careful validation and transparency in modelling processes.

### Limitations

Our study has several limitations. First, MPoRT relies on self-reported data, susceptible to recall bias and measurement error, particularly for variables like alcohol consumption, physical activity, and diet. Although some important risk factors like smoking include historical exposure (e.g., former smoking status), the accuracy of self-reported information can vary and affect risk estimation, but there are approaches to assess, characterize, and adjust for these concerns.[53,54]

Second, MPoRT was developed and validated using data from Canada and the United States, two high-income countries with cultural similarities. The model’s performance may differ in middle or low-income countries, where death from infectious diseases, injuries and other causes are more prevalent than in Canada and the United States. Nonetheless, death has become more prevalent worldwide for conditions related to risk exposures using MPoRT, and we anticipate these same risk exposures are discriminating for mortality risk in other settings.[24] However, MPoRT requires an assessment before application in these settings.

Third, we acknowledge the potential for unmeasured confounding variables, such as mental health status, psychosocial stress, environmental exposures, and social support, which were not included in the model but can influence mortality risk. [55]

### Bias, fairness, and ethical considerations

Predictive algorithms carry inherent risks of bias, potentially perpetuating health inequities if certain populations are inadequately represented. MPoRT, however, has several strengths that enhance its fairness and representativeness. The algorithm includes a wide range of sociodemographic predictors—more extensive than many existing risk models—allowing detailed mortality risk assessment across diverse population subgroups. Additionally, community-based survey data, broadly representative of general populations in Canada and the United States, provides greater external validity than clinical or administrative datasets typically used in similar studies.

Nonetheless, some groups remain underrepresented due to sampling exclusions, including residents of remote areas, institutionalized individuals, military personnel, and Indigenous communities. Such exclusions may introduce bias, potentially limiting MPoRT’s accuracy and fairness in these populations.

The reliance on self-reported predictors might also differentially affect risk estimation accuracy across socioeconomic or cultural groups, as health perceptions and reporting can vary systematically. Thus, careful validation and recalibration remain essential before applying MPoRT globally or in disadvantaged communities.

Ethically, predictive algorithms like MPoRT must be transparently communicated to users and decision-makers, indicating uncertainty in predictions and potential implications for resource allocation and intervention prioritization.

## Conclusions

All-cause mortality risk in the general population can be discriminately and accurately assessed using information focusing on sociodemographic, behavioural and chronic disease status. Model discrimination was maintained in external validation in two countries (Canada and the United States). Recalibration for different countries appears feasible, given the widespread availability of national mortality statistics.

Personalized decision-making for multiple chronic disease prevention and management is facilitated by estimating how different prevention interventions will extend life expectancy (years of life lost). Whether these metrics improve decision-making requires further assessment in the clinic setting.

## Data Availability

Three sources of data were used for this study:
Ontario CCHS (ICES Data): The dataset from this study is held securely in coded form at ICES. While data-sharing agreements prohibit ICES from making the dataset publicly available, access may be granted to those who meet pre-specified criteria for confidential access (available at www.ices.on.ca/DAS). The full dataset creation plan and underlying analytic code are available from the authors upon reasonable request.
United States NHIS Data: The National Health Interview Survey (NHIS) public-use data files are freely available to researchers and the general public without special permissions or restricted access, and can be downloaded directly from the NCHS website (https://www.cdc.gov/nchs/nhis/data-questionnaires-documentation.htm). Restricted-use data files are available through the NCHS Research Data Center (RDC) subject to an application process.
National Canadian CCHS Data: The Canadian version of the CCHS linked to mortality is available at Statistics Canada Regional Data Centres (RDCs). Access to these secure microdata files is restricted to affiliated researchers who apply and are approved through Statistics Canada (https://www.statcan.gc.ca/en/microdata/data-centres).

## Acknowledgements

Contributors

DM conceived the study and developed the design in consultation with the other authors. The analysis was completed by AE, PF, SF, MT, JL, JS, and DH. All authors contributed to interpreting the data. DM and CB drafted the manuscript, and all authors contributed substantially to its revision. All authors approved the final version to be published and agreed to be accountable for all aspects of the work.

## Disclosure and Disclaimer

This study was supported by ICES, which is funded by an annual grant from the Ontario Ministry of Health and Long-Term Care (MOHLTC). The opinions, results and conclusions reported in this paper are those of the authors and are independent from the funding sources. No endorsement by ICES, the Ontario MOHLTC or Statistics Canada is intended or should be inferred. Parts of this material are based on data and/or information compiled and provided by CIHI. However, the analyses, conclusions, opinions and statements expressed in the material art those of the authors, and not necessarily those of CIHI.

The use of data in this project was authorized under section 45 of Ontario’s Personal Health Information Protection Act, which does not require review by a Research Ethics Board.

## Data Access

Three sources of data were used for this study.

Ontario sample of the Canadian Community Health Survey, used for the original derivation algorithm, was linked using unique encoded identifiers and analyzed at ICES. The dataset from this study is held securely in coded form at ICES. While data-sharing agreements prohibit ICES from making the dataset publicly available, access may be granted to those who meet pre-specified criteria for confidential access, available at www.ices.on.ca/DAS. The full dataset creation plan and underlying analytic code are available from the authors upon request, understanding that the programs may rely upon coding templates or macros unique to ICES.

The United States NHIS used to validate the MPoRT algorithm and generate a harmonized Canadian-US algorithm. The NHIS was conducted by the National Center for Health Statistics (NCHS). The NHIS public-use data files are freely available to researchers and the general public without special permissions or restricted access. These files can be downloaded directly from the NCHS website (https://www.cdc.gov/nchs/nhis/data-questionnaires-documentation.htm). The public-use files contain de-identified data to protect respondent confidentiality while still providing a wealth of information for research purposes. For researchers requiring access to more detailed data not available in the public-use files, the NCHS Research Data Center (RDC) offers access to restricted-use data files, subject to an application process and adherence to strict confidentiality protocols. Further information about both public-use and restricted-use data access, documentation, and user guidelines can be found on the NCHS website.https://www.cdc.gov/nchs/nhis/index.htm

The Canadian version of CCHS linked to mortality is available at Statistics Canada Regional Data Centres, administered by Statistics Canada. Access to the data is restricted to researchers affiliated with academic, government, or other recognized institutions. To access the CCHS microdata, researchers must apply for access through one of Statistics Canada’s Research Data Centres (RDCs) across Canada. These secure facilities provide access to confidential data by the confidentiality provisions of the Statistics Act. Further information about access procedures, eligibility, and application requirements can be found on the Statistics Canada website (https://www.statcan.gc.ca/en/microdata/data-centres).

## Additional files

### Additional file 1. Predictor variables for the Mortality Population Risk Tool (MPoRT) for Canada and United States national data

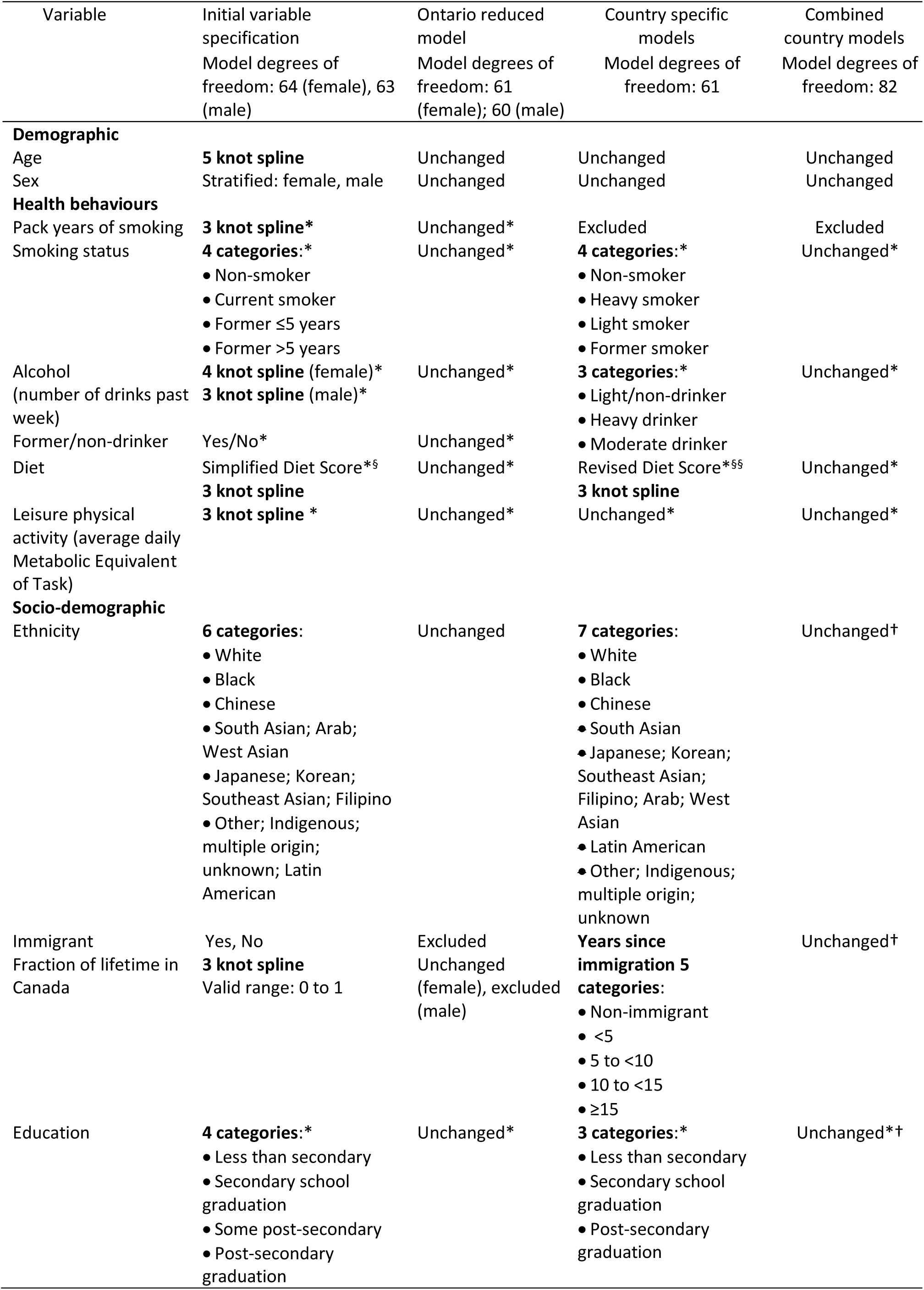
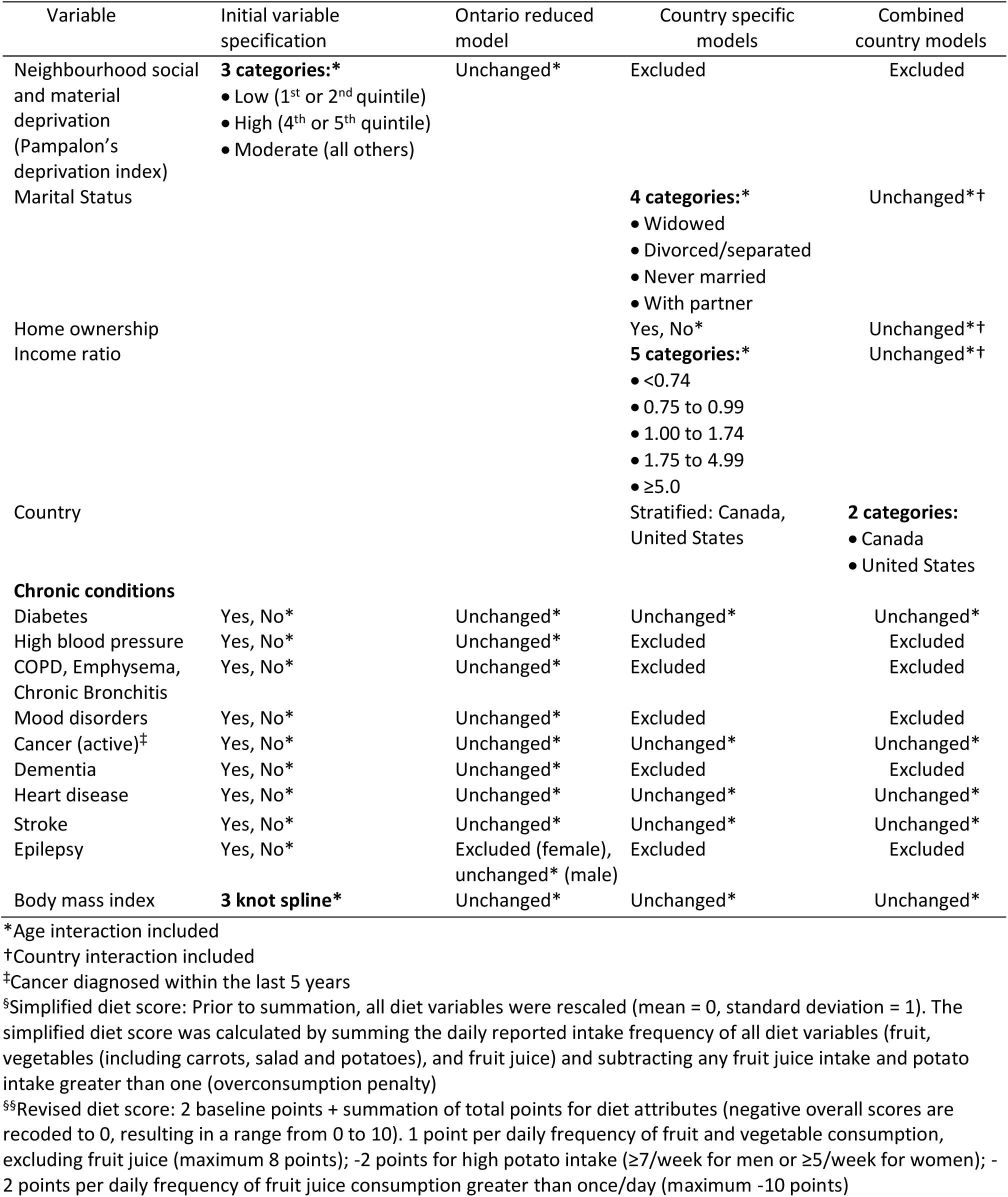

### Additional file 2. Detailed methods

#### Identification of predictors

For the derivation model, in addition to sex (stratified), 22 predictor variables were identified and selected based on previous mortality and cardiovascular disease risk algorithms derived using the same data source.(8–10) An indicator variable for immigration status together with fraction of life lived in Canada was used to account for both recent and non-recent immigrants. Indicator variables for smoking status were created to allow inclusion of smoking pack-years as a continuous predictor. Detailed definitions and measurement of these variables are presented in Table 1. The specific wording of questions is available within the survey documentation.

#### Data cleaning and coding of predictors

Data cleaning and coding was completed without examining outcome-risk factor associations. After inspection of histograms and boxplots, continuous variables were truncated to the 99.5^th^ percentile.

#### Missing data

We used multiple imputation to impute missing values on predictor variables.(3) The imputation model consisted of the full list of predictor variables, time to event and censoring variables, and auxiliary variables—that is, variables that are not predictors but may nevertheless be useful in generating imputed values (for example, income and self-perceived health). We generated five multiple imputation datasets. The imputation procedure employed predictive mean matching to generate imputed values and used the bootstrap to approximate the process of drawing predicted values from a full Bayesian predictive distribution. Continuous variables in the imputation model were transformed using restricted cubic splines. The final prediction model was estimated separately for each imputation-completed dataset and the results combined using the rules developed by Rubin(11) to account for imputation uncertainty.

#### Model specification

We fit a preliminary model that included an initial degree of freedom allocation for each predictor and age interaction for all but three predictors (immigrant status, fraction of time in Canada, and ethnicity). Continuous predictors were flexibly modelled using restricted cubic splines, i.e., piecewise cubic functions that are smooth at the knots and restricted to be linear in the tails. The knots were placed at fixed quantiles of the distribution: in particular, at the 5^th^, 27.5^th^, 50^th^, 72.5^th^ and 95^th^ percentiles for 5 knot splines; at the 5^th^, 35^th^, 65^th^ and 95^th^ percentiles for 4 knot splines; and, at the 10^th^, 50^th^ and 90^th^ percentiles for 3 knot splines.

The initial model specification, presented in **Table 1**, included 64 degrees of freedom (41 main, 23 interaction) for the female model and 63 degrees of freedom (40 main, 23 interaction), compared to a possible maximum (based on number of events) of 734 (female) and 796 (male). We allocated the final degrees of freedom to individual predictors based on a partial test of association with the outcome. Partial association chi-squared statistics for each predictor variable minus their degrees of freedom were plotted in descending order. We retained the initial degrees of freedom for variables with higher predictive potential, but predictors with lower predictive potential were modelled with reduced degrees of freedom, i.e., as simple linear terms or after combining infrequent categories. Interaction terms, specified above, were added to the final model and were restricted to linear terms.

The full models were specified in accordance with the pre-specified plan of maintaining all predictors in the model but with reduced DFs, reflecting the importance in the partial correlation plots. After applying the step-down procedure, the final application (reduced) model had 61 and 60 degrees of freedom with 20 predictors and 17 and 18 interaction terms for female and male models, respectively.

#### Model estimation

Models were estimated using a proportional hazards model. All predictors were centred about their means prior to analysis. The proportionality assumption was assessed using plots of raw and smoothed scaled Schoenfeld residuals versus time for each predictor. Survey weights were not used for model development. We recommend survey weight for population application when available or used in population application data. Analyses were conducted using coxph and Harrell’s HMisc(3) and rms packages of functions in R(12) and SAS v9.3.

#### Assessment of model performance

Nagelkerke’s *R*^2^ and the Brier score were calculated as overall accuracy measures. Discrimination was assessed using Harrell’s overall concordance statistic, with 95% confidence intervals estimated using bootstrap samples.

Calibration plots were created by comparing mean 5-year predicted probabilities with cumulative incidence function estimates of observed rates stratified by deciles of predicted risk. The calibration slope was estimated by regressing the observed risk (1-survival) on the mean predicted risk within predicted risk groups (20 groups). Deviation from a slope of 1 was tested using a Wald test. The calibration slope reflects the combined effect of over-fitting to the derivation data and true differences in effects of predictors. All model performance measures were calculated using the first of the multiply imputed datasets.

No major violation of the proportional hazards assumption was observed.

#### Risk groups

Subgroup validation was implemented as a conceptually easy calibration check by comparing observed and predicted risks within predefined subgroups of importance to clinicians and policymakers.(10) We examined subgroups using predefined criteria for clinically or policy-relevant calibration standards (<20% difference between observed and predicted estimates for categories with prevalence higher than 5%). In total, 124 subgroups were defined based on age, behavioural risk exposure categories, health regions, socio-demographic groups, hypertension status, and diabetes status.

#### Estimation of the final parsimonious model

The parsimonious model was generated by deleting variables to a desired degree of accuracy based on contribution to model *R*^2^.(13) To maximize the amount of data for the final model, the final regression coefficients were estimated using the combined data from both the derivation and validation cohorts with outcome events updated to reflect the most recent years available.

### Additional file 3. Study flow Ontario derivation and validation cohorts

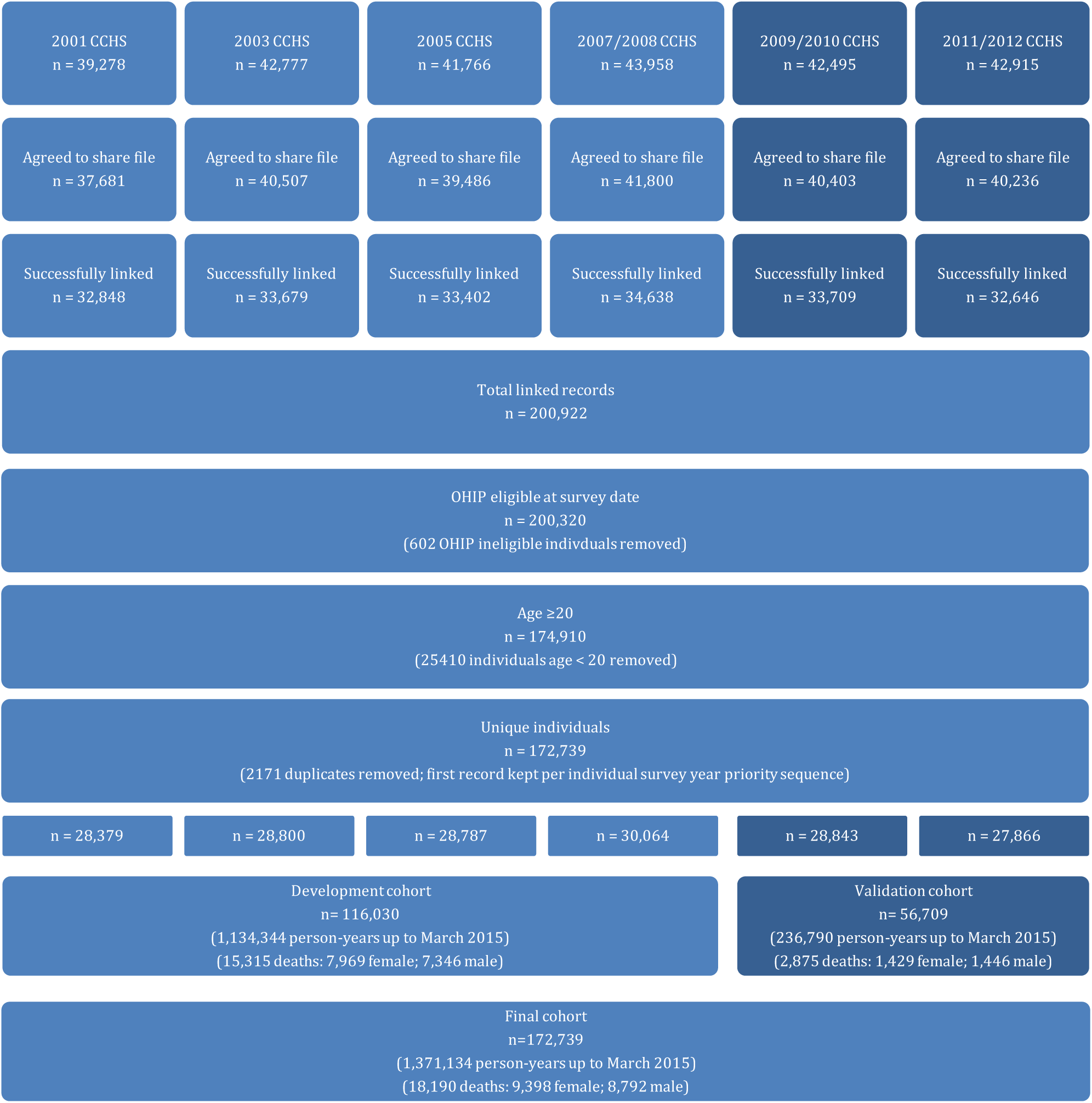

### Additional file 4. Number of deaths, person-years follow up, and death rate per 1000 person-years of observation in derivation and validation cohorts

|  | Ontario derivation cohort |  |  | Ontario validation cohort |  |  | United States validation cohort |  |  | Canada validation cohort |  |  |
| --- | --- | --- | --- | --- | --- | --- | --- | --- | --- | --- | --- | --- |
|  | Number of deaths | Person-years | Death rate per 1000 person-years (95% CI) | Number of deaths | Person-years | Death rate per 1000 person-years (95% CI) | Number of deaths | Person-years | Death rate per 1000 person-years (95% CI) | Number of deaths | Person-years | Death rate per 1000 person-years (95% CI) |
| Females |  |  |  |  |  |  |  |  |  |  |  |  |
| [20 – 30) | 35 | 87,280 | 0.4 (0.3-0.6) | 9 | 17,407 | 0.5 (0.2-1.0) | 27 | 98,466 | 0.3 (0.2-0.4) | 18 | 27,578 | 0.7 (0.4-1.0) |
| [30 – 40) | 118 | 119,025 | 1.0 (0.8-1.2) | 13 | 19,262 | 0.7 (0.4-1.2) | 96 | 173,002 | 0.6 (0.4-0.7) | 54 | 57,037 | 0.9 (0.7-1.2) |
| [40 – 50) | 298 | 112,664 | 2.7 (2.4-3.0) | 32 | 18,105 | 1.8 (1.2-2.5) | 228 | 166,787 | 1.4 (1.2-1.5) | 167 | 60,029 | 2.8 (2.4-3.2) |
| [50 – 60) | 660 | 111,190 | 5.9 (5.5-6.4) | 93 | 24,158 | 3.9 (3.1-4.7) | 635 | 180,532 | 3.5 (3.2-3.8) | 269 | 51,404 | 5.2 (4.6-5.9) |
| [60 – 70) | 1,346 | 91,619 | 14.7 (13.9-15.5) | 233 | 25,109 | 9.3 (8.1-10.6) | 1,298 | 161,672 | 8.0(7.6-8.5) | 401 | 36,217 | 11.1 (10.0-12.1) |
| [70 – 80) | 2,696 | 72,053 | 37.4 (36.0-38.9) | 412 | 17,370 | 23.7 (21.5-26.1) | 2,288 | 124,391 | 18.4 (17.6-19.1) | 713 | 27,840 | 25.6 (23.8-27.5) |
| [80+) | - | - | - | - | - | - | 4,980 | 89,578 | 55.6 (54.1-57.1) | 1,746 | 22,769 | 76.7 (73.2-80.1) |
| [80 – 90) | 2,464 | 28,784 | 85.6 (82.3-89.1) | 505 | 9,914 | 50.9 (46.6-55.6) | - | - | - | - | - | - |
| [90 – 105] | 352 | 1,977 | 178.0 (159.9-197.6) | 132 | 972 | 135.8 (113.6-161.0) | - | - | - | - | - | - |
| Overall | 7,969 | 624,592 | 12.8 (12.5-13.0) | 1,429 | 132,298 | 10.8 (10.3-11.4) | 9,552 | 994,431 | 9.6 (9.4-9.8) | 3,368 | 282,878 | 11.9 (11.5-12.3) |
| Males |  |  |  |  |  |  |  |  |  |  |  |  |
| [20 – 30) | 43 | 69,744 | 0.6 (0.5-0.8) | 9 | 15,119 | 0.6 (0.3-1.1) | 60 | 80,730 | 0.7 (0.6-0.9) | 28 | 22,243 | 1.3 (0.8-1.7) |
| [30 – 40) | 140 | 103,738 | 1.4 (1.1-1.6) | 10 | 15,414 | 0.7 (0.3-1.2) | 118 | 146,114 | 0.8 (0.7-1.0) | 68 | 45,013 | 1.5 (1.2-1.9) |
| [40 – 50) | 389 | 107,367 | 3.6 (3.3-4.0) | 47 | 16,495 | 2.9 (2.1-3.8) | 342 | 161,932 | 2.1 (1.9-2.3) | 154 | 48,608 | 3.2 (2.7-3.7) |
| [50 – 60) | 816 | 93,913 | 8.7 (8.1-9.3) | 140 | 19,400 | 7.2 (6.1-8.5) | 864 | 154,268 | 5.6 (5.2-6.0) | 378 | 41,424 | 9.1 (8.2-10.0) |
| [60 – 70) | 1,611 | 74,125 | 21.7 (20.7-22.8) | 284 | 19,958 | 14.2 (12.6-16.0) | 1,654 | 136,623 | 12.1 (11.5-12.7) | 506 | 28,422 | 17.8 (16.3-19.3) |
| [70 – 80) | 2,630 | 47,503 | 55.4 (53.3-57.5) | 409 | 12,481 | 32.8 (29.7-36.1) | 2,898 | 93,330 | 31.1 (29.9-32.2) | 748 | 18,845 | 39.7 (36.9-42.5) |
| [80+) | - | - | - | - | - | - | 3739 | 46,457 | 80.5 (78.0-83.0) | 1091 | 10,477 | 104.1 (98.3-110.0) |
| [80 – 90) | 1,572 | 12,717 | 123.6 (117.6-129.9) | 458 | 5,256 | 87.1 (79.3-95.5) | - | - | - | - | - | - |
| [90 – 105] | 145 | 645 | 224.9 (189.8-264.6) | 89 | 369 | 241.3 (193.8-296.9) | - | - | - | - | - | - |
| Overall | 7,346 | 509,752 | 14.4 (14.1-14.7) | 1,446 | 104,492 | 13.8 (13.1-14.6) | 9675 | 819,453 | 11.8 (11.6-12.0) | 2973 | 215,032 | 13.8 (13.3-14.3) |

### Additional file 5. Population characteristics of the Mortality Population Risk Tool (MPoRT) Ontario derivation and validation cohorts

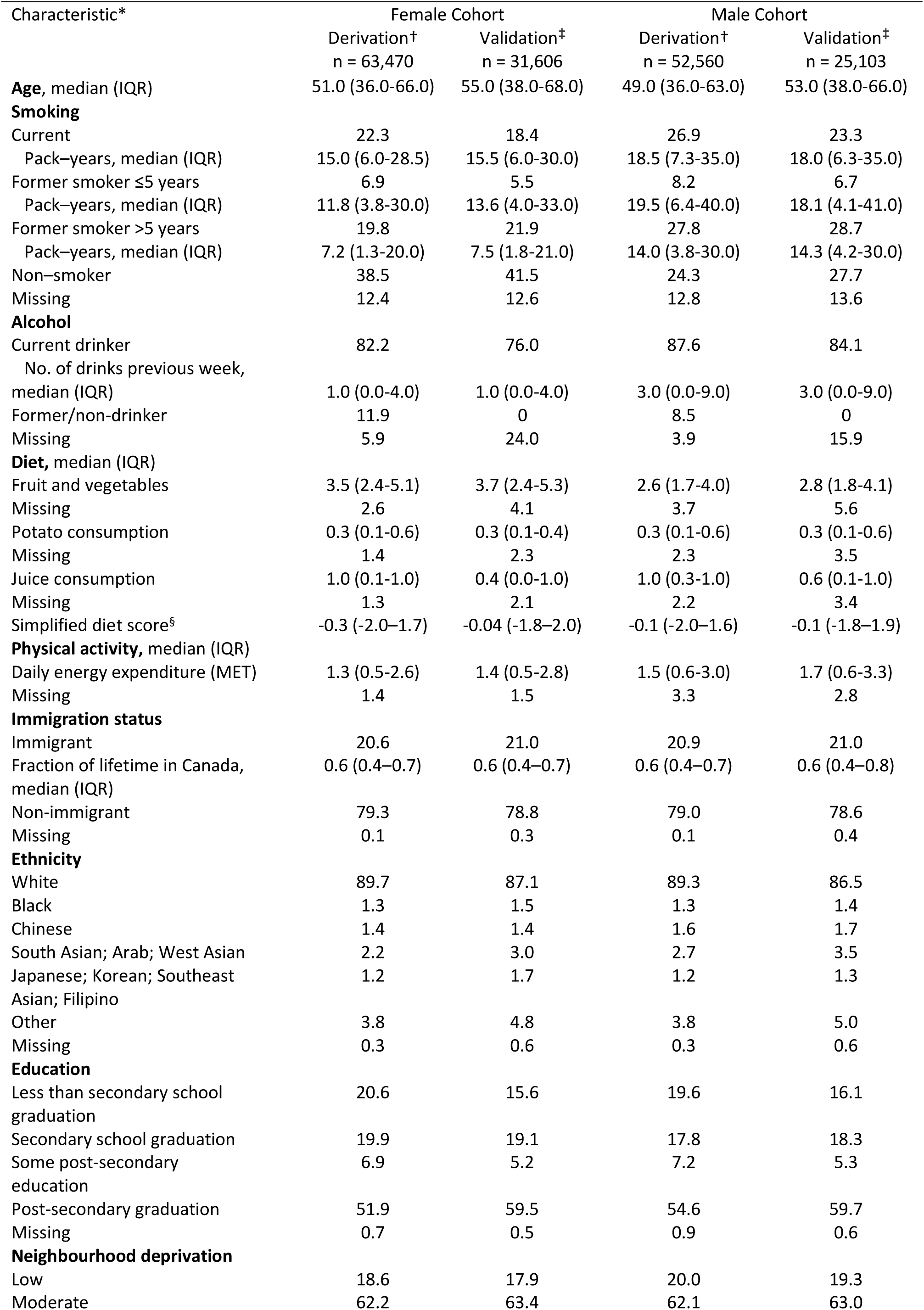
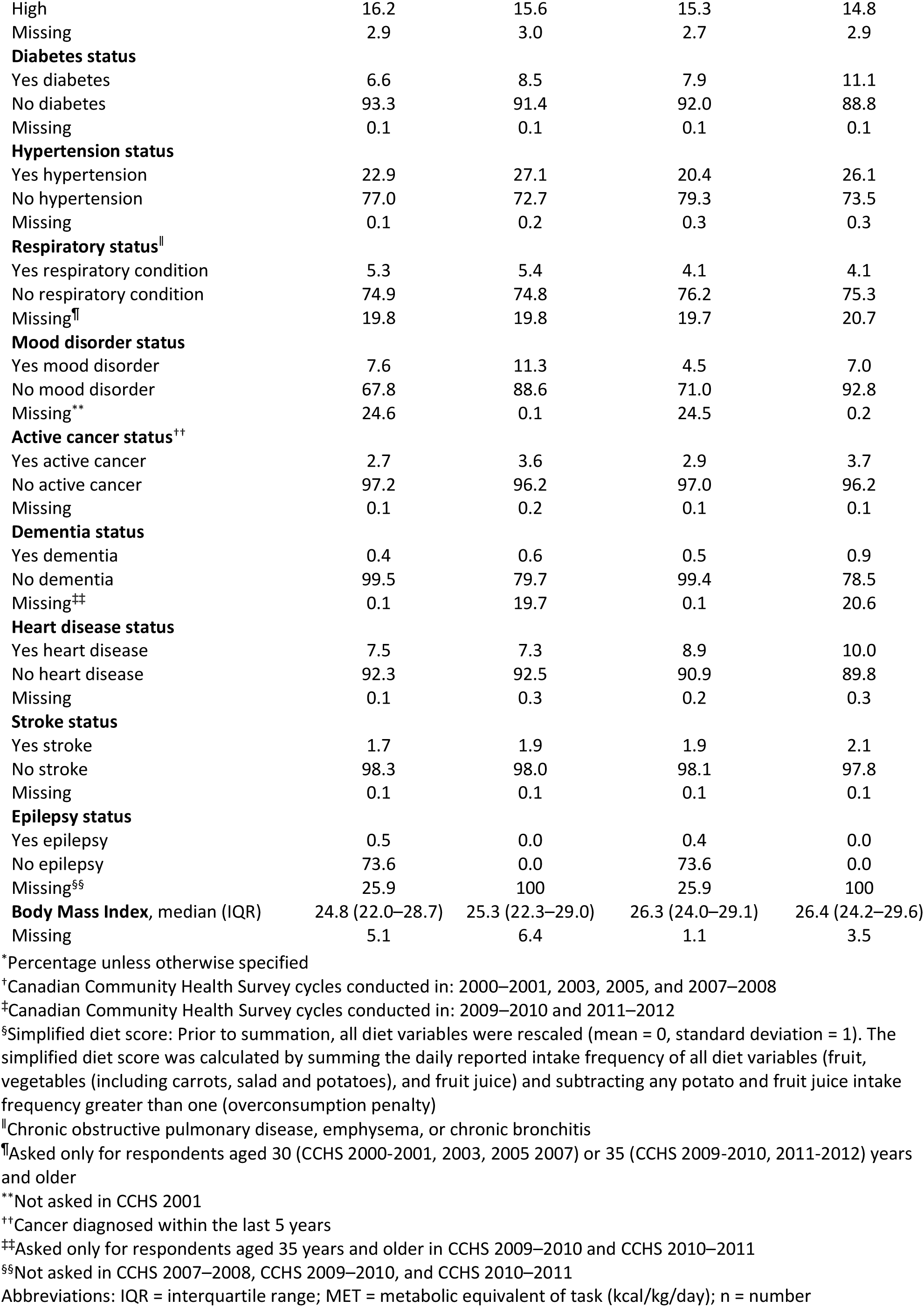

### Additional file 6. Population characteristics of national Canadian Community Health Survey (CCHS) and National Health Interview Survey (NHIS) external validation cohorts

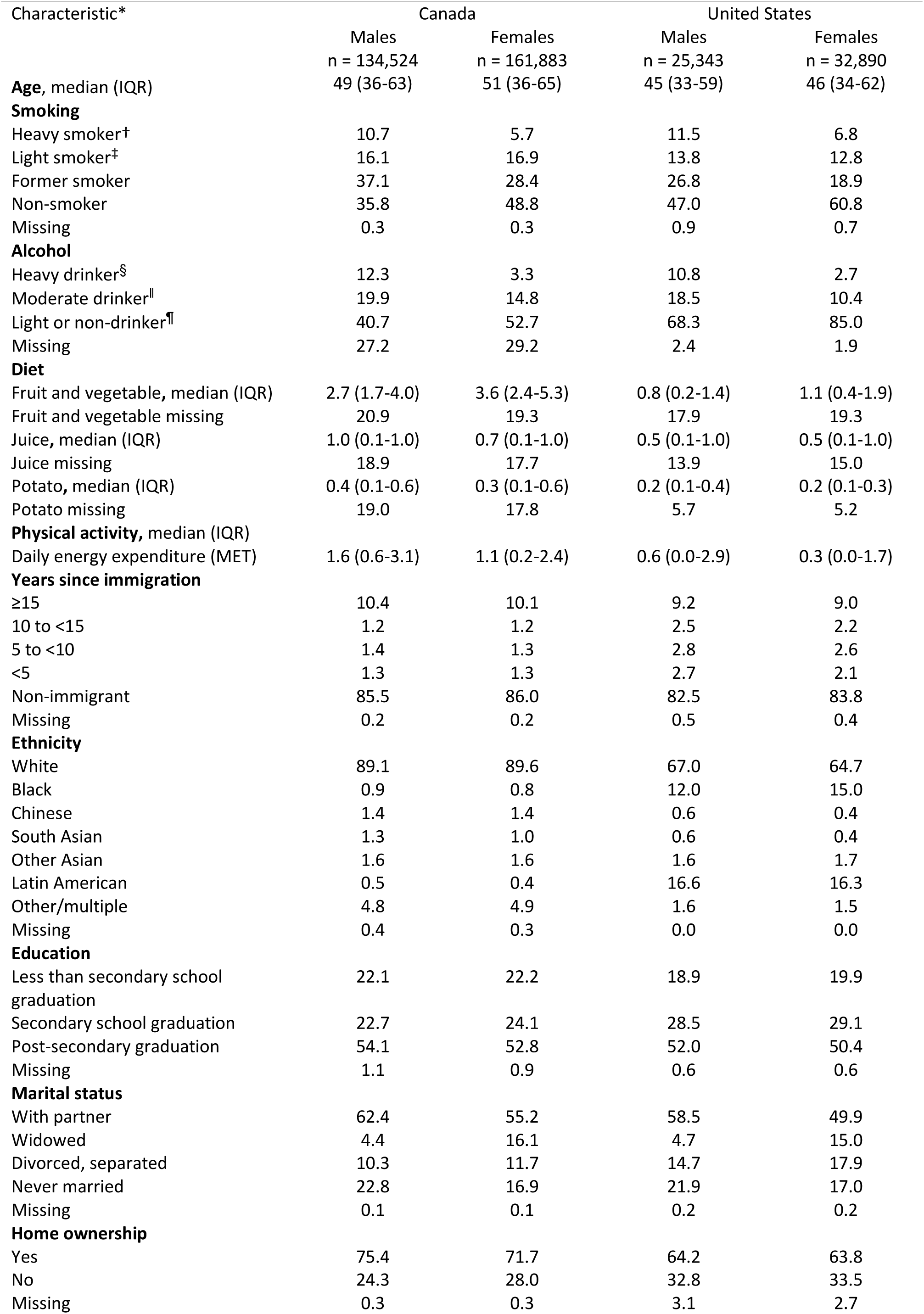
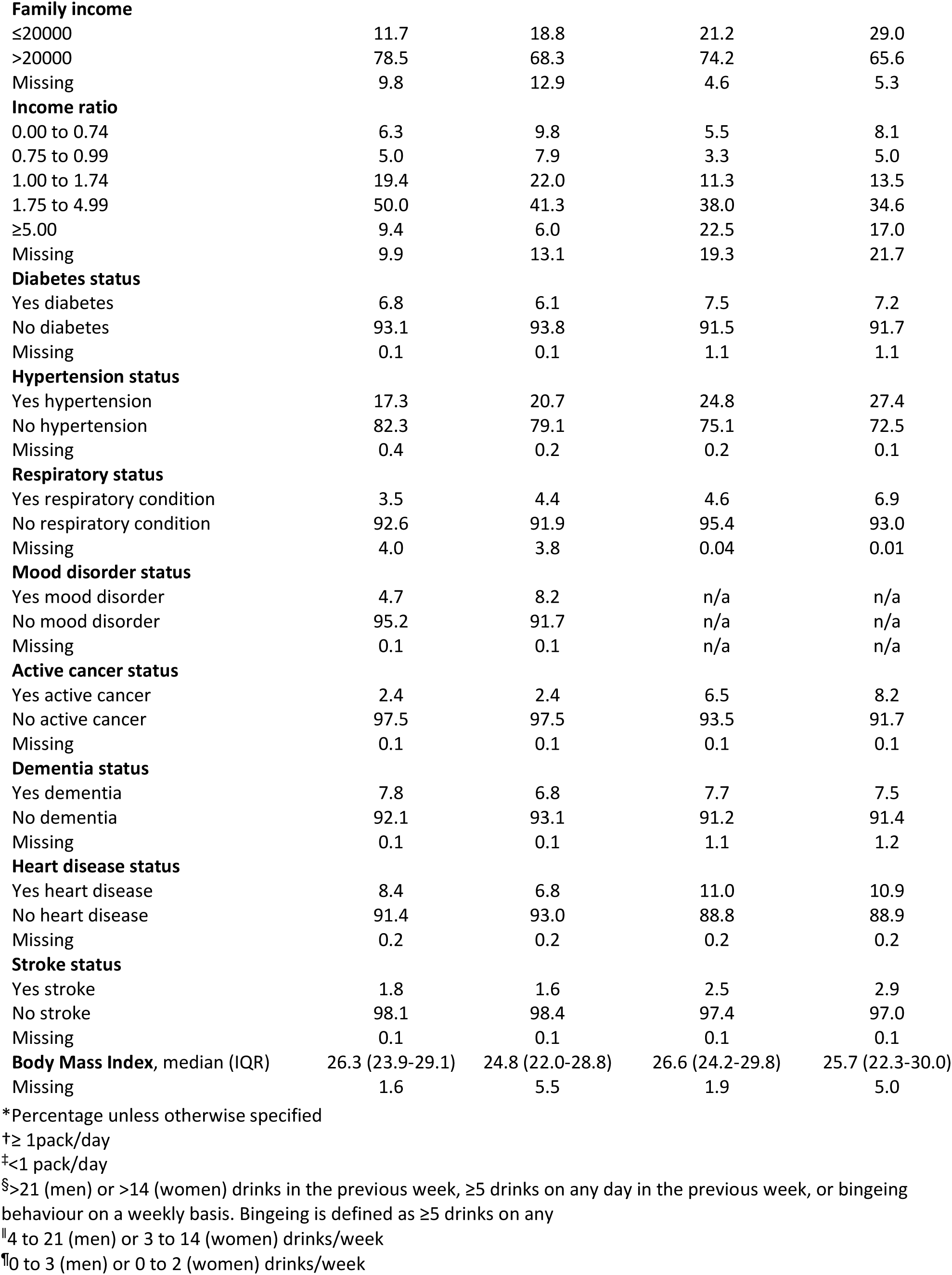

### Additional file 7. MPoRT beta values

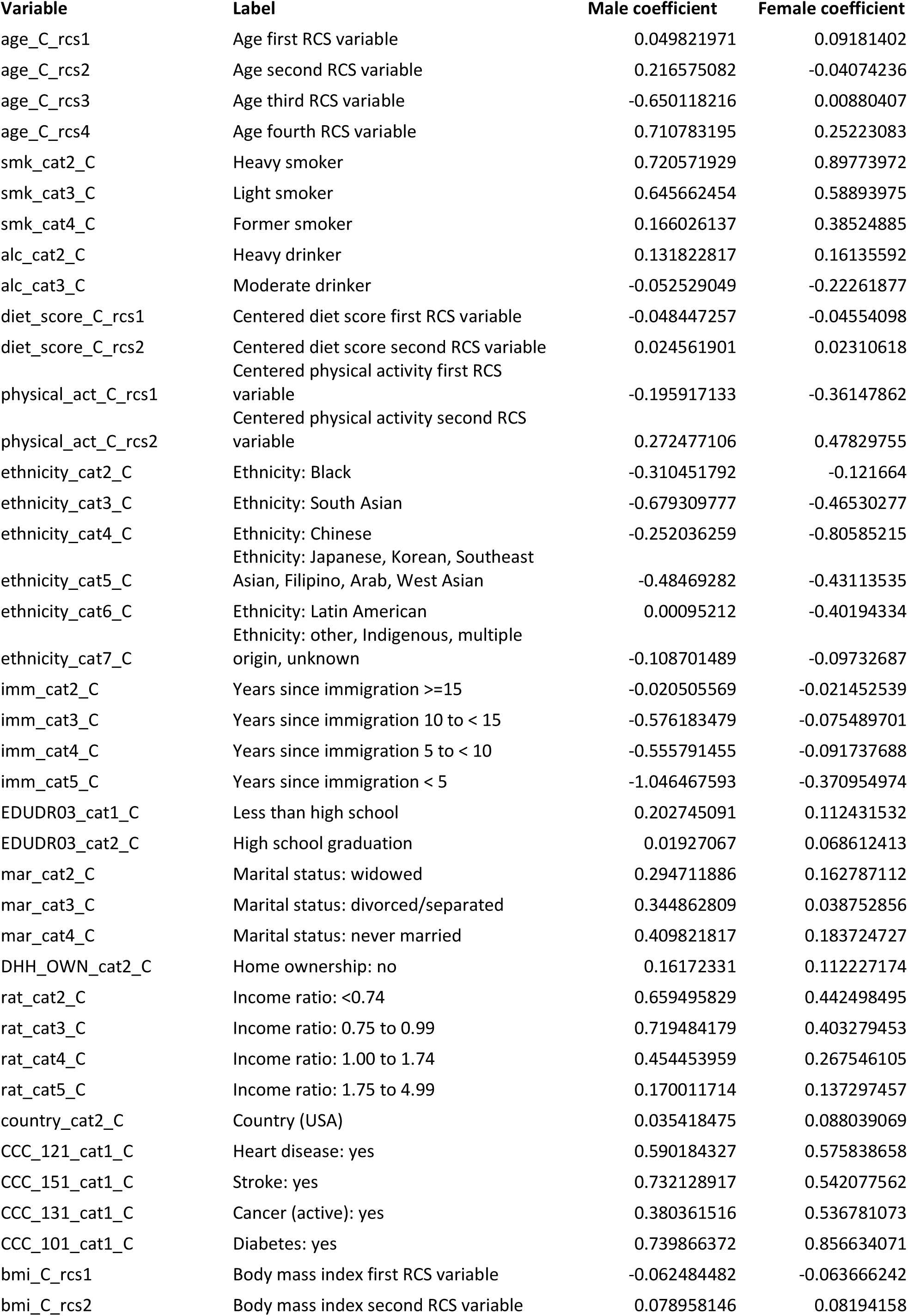
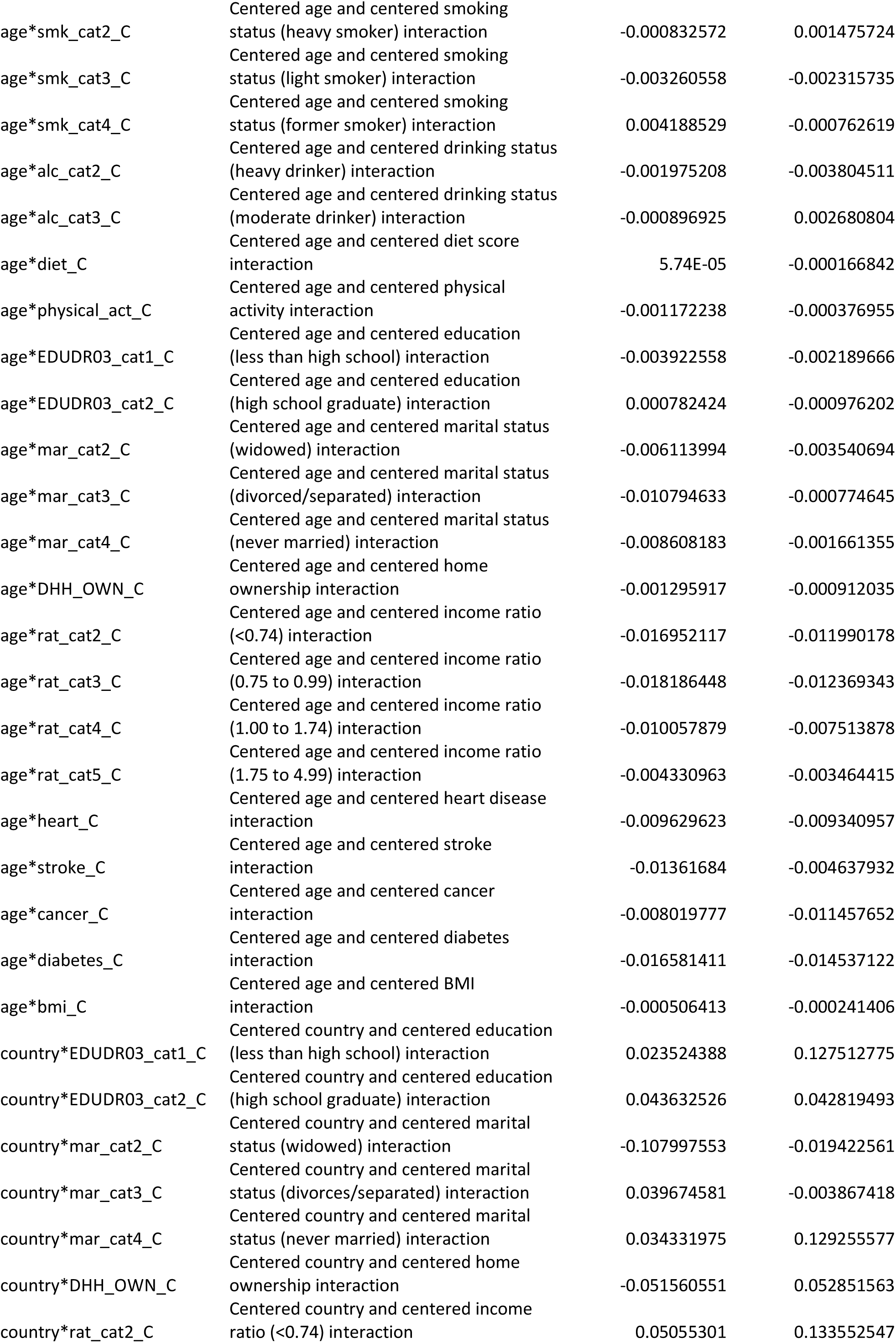
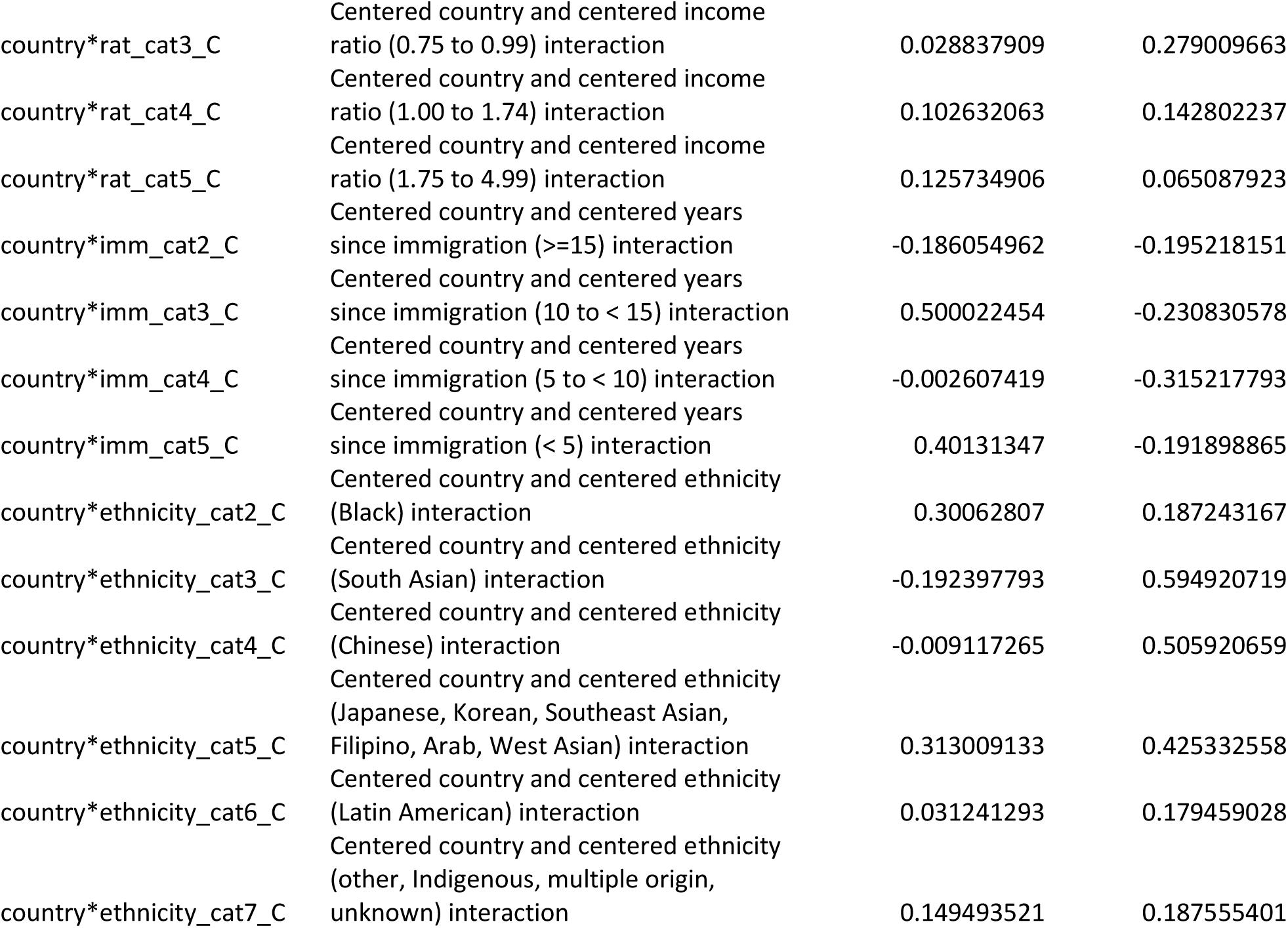

### Additional file 8. MPoRT formula for risk

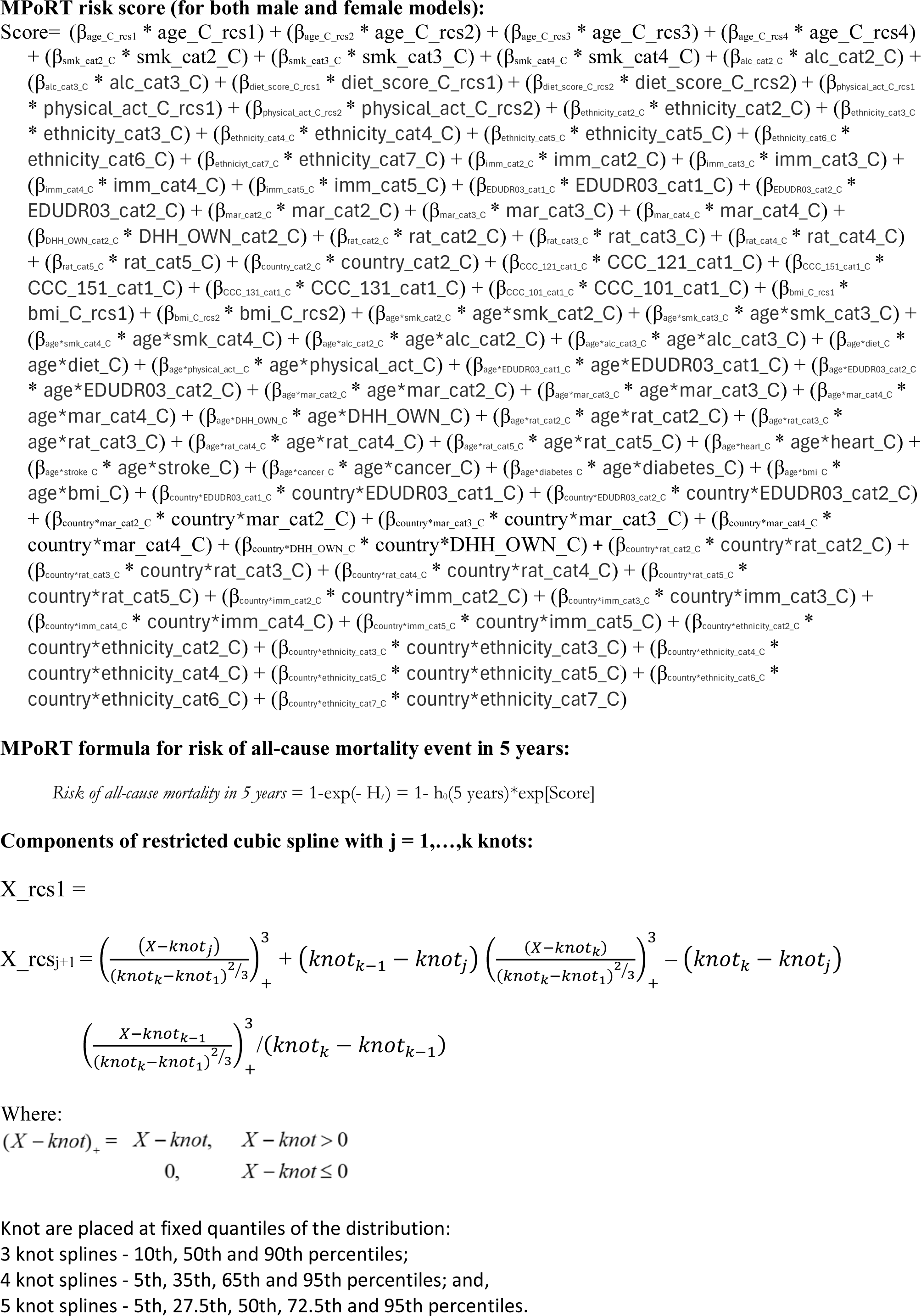

### Additional file 9. Observed versus predicted by subgroups – Ontario validation cohorts

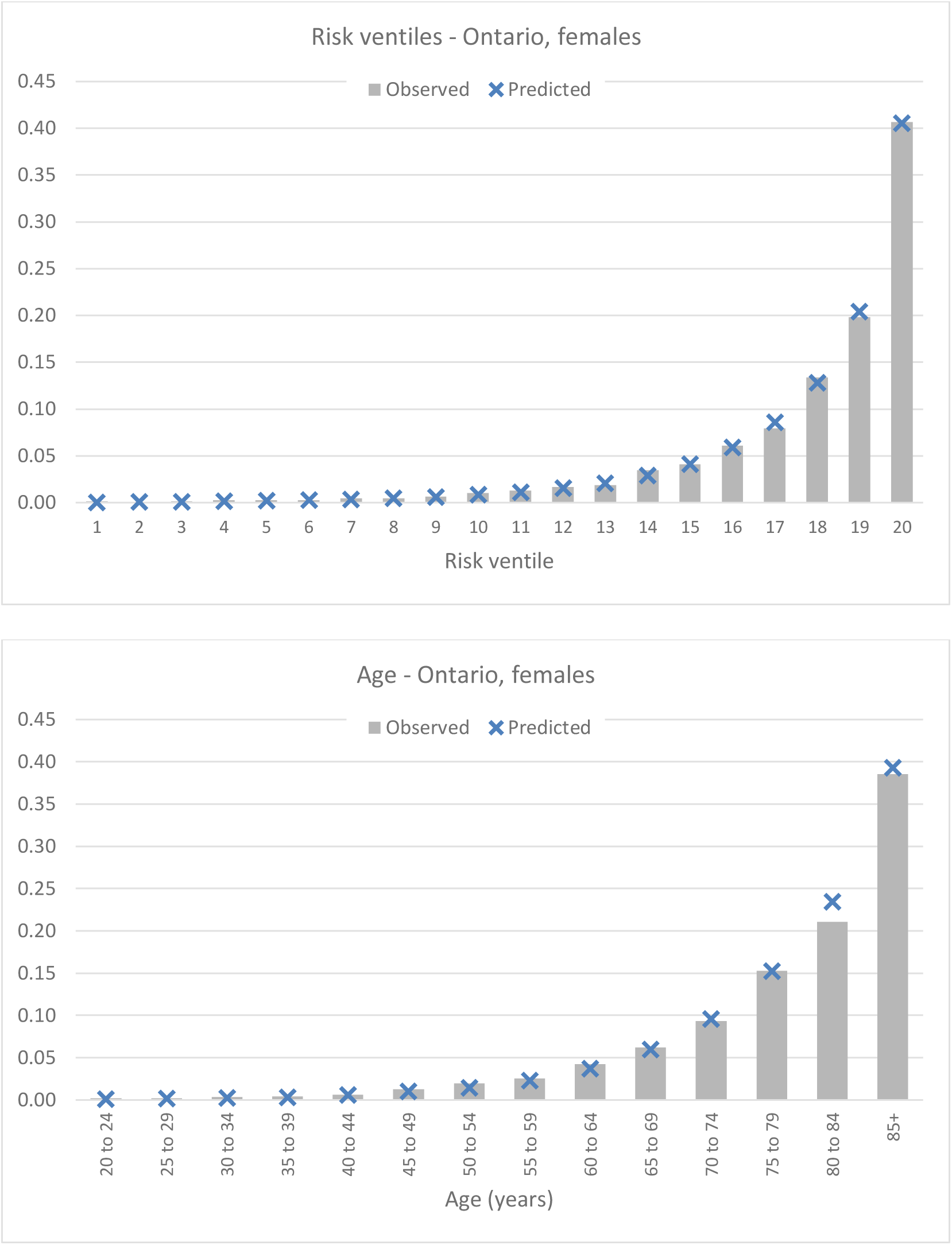
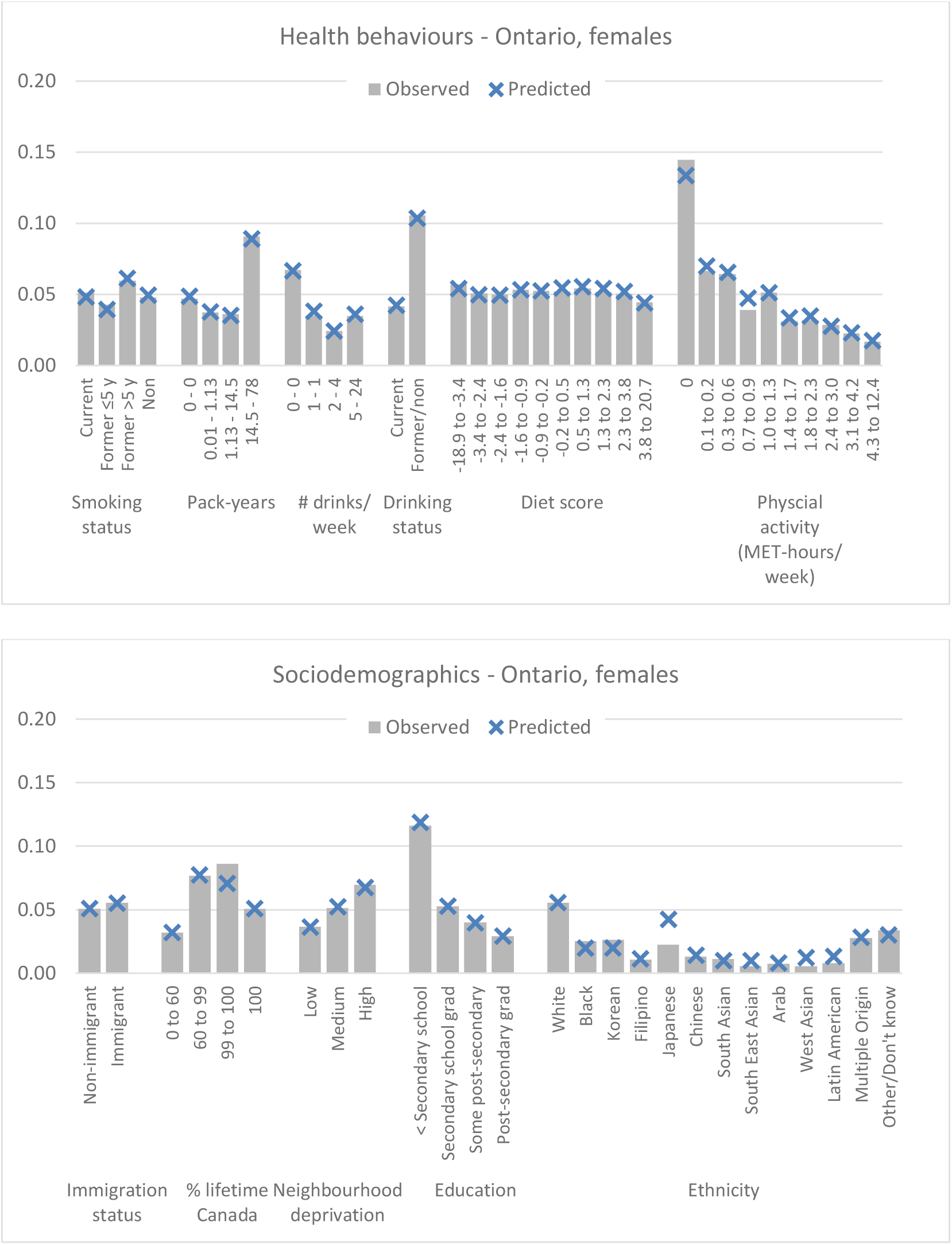
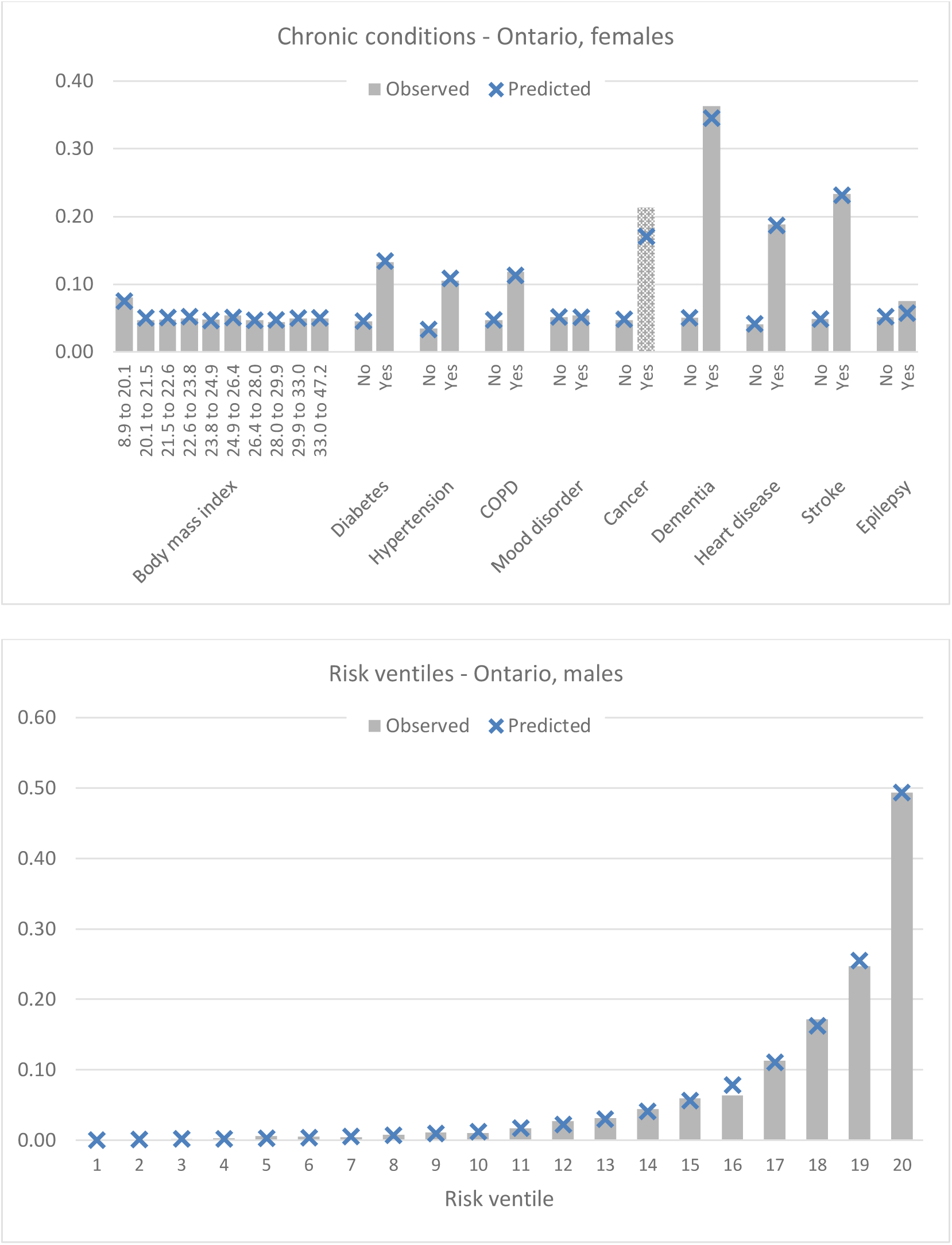
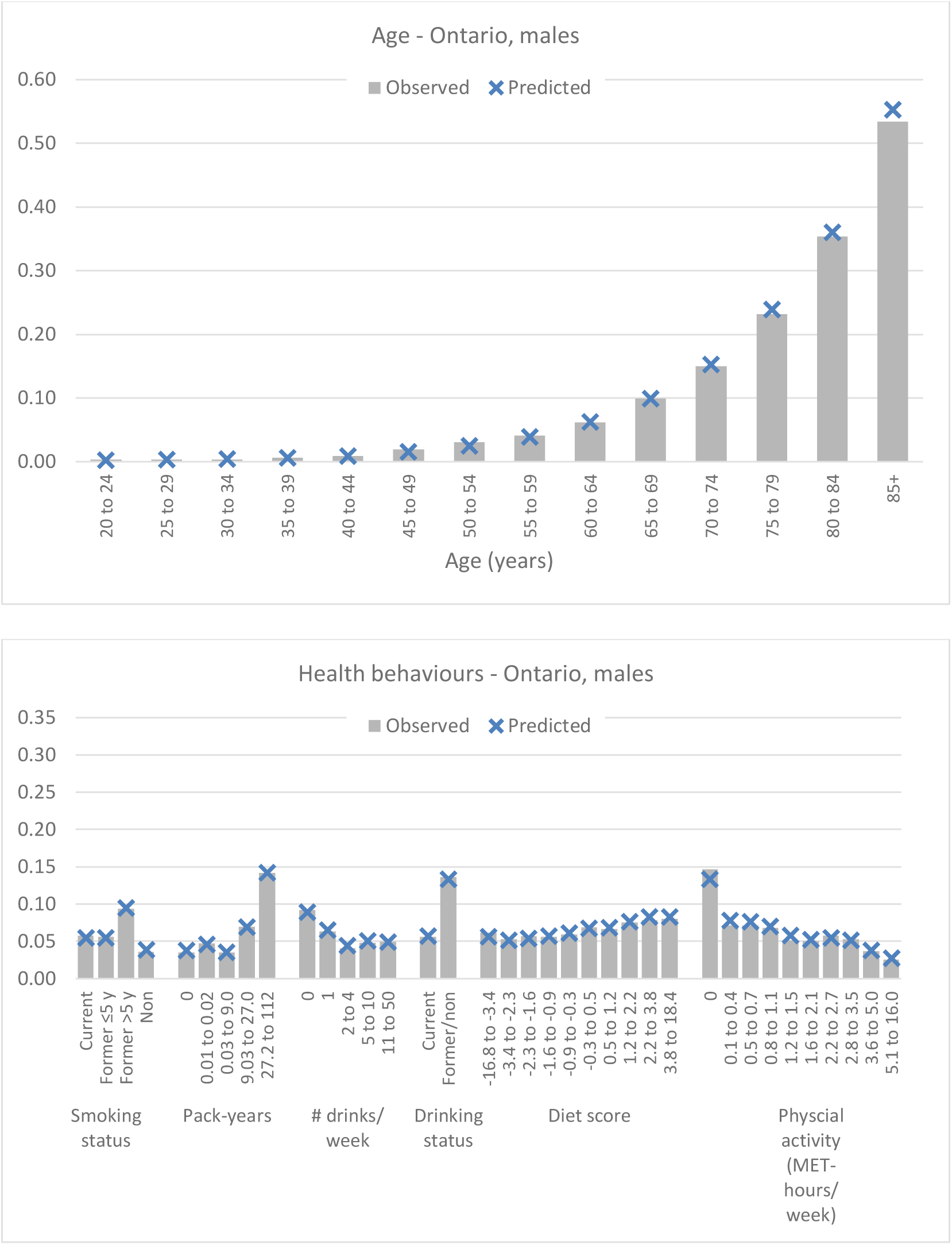
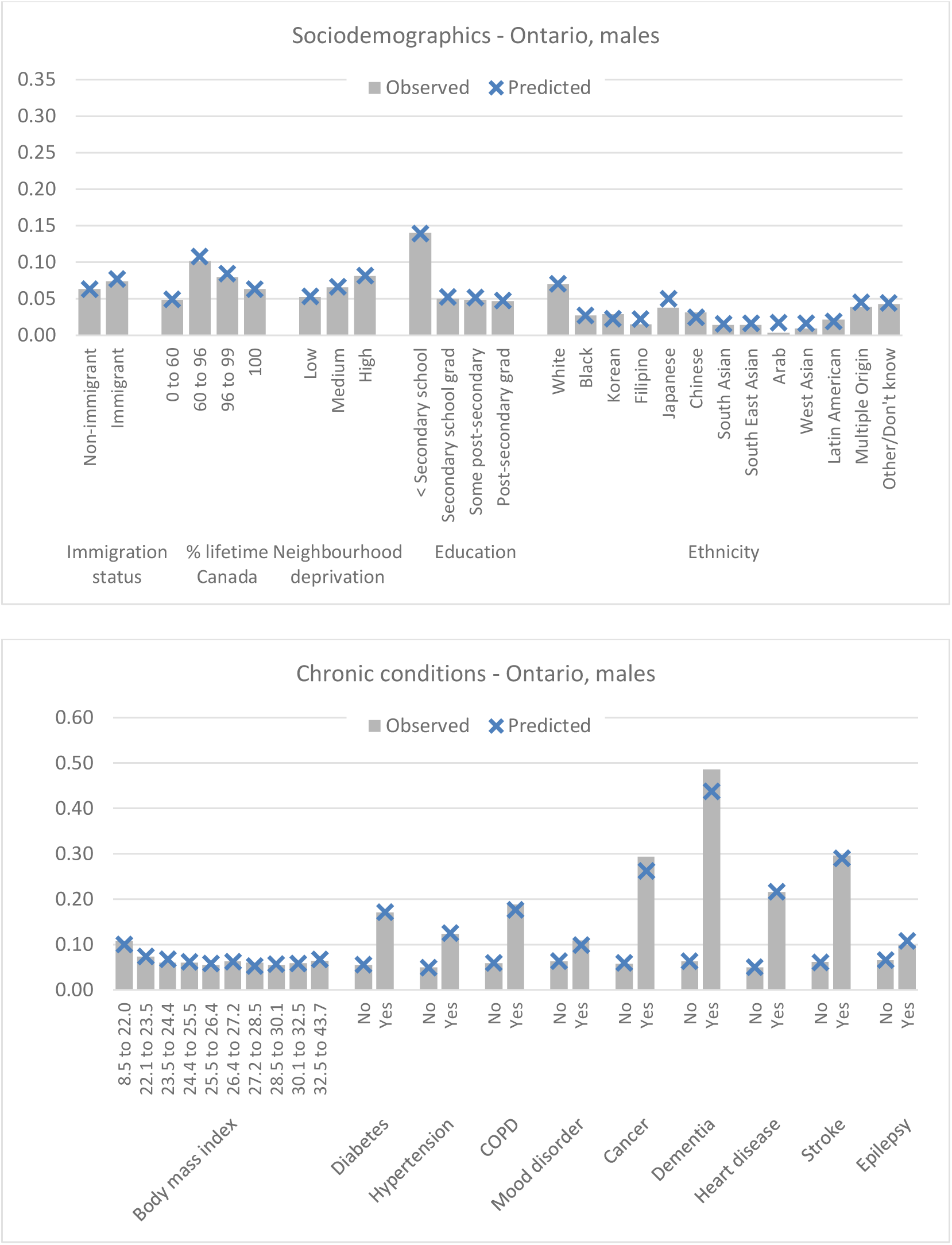

### Additional file 10. Observed versus predicted by subgroups – females external validation, US and Canada combined

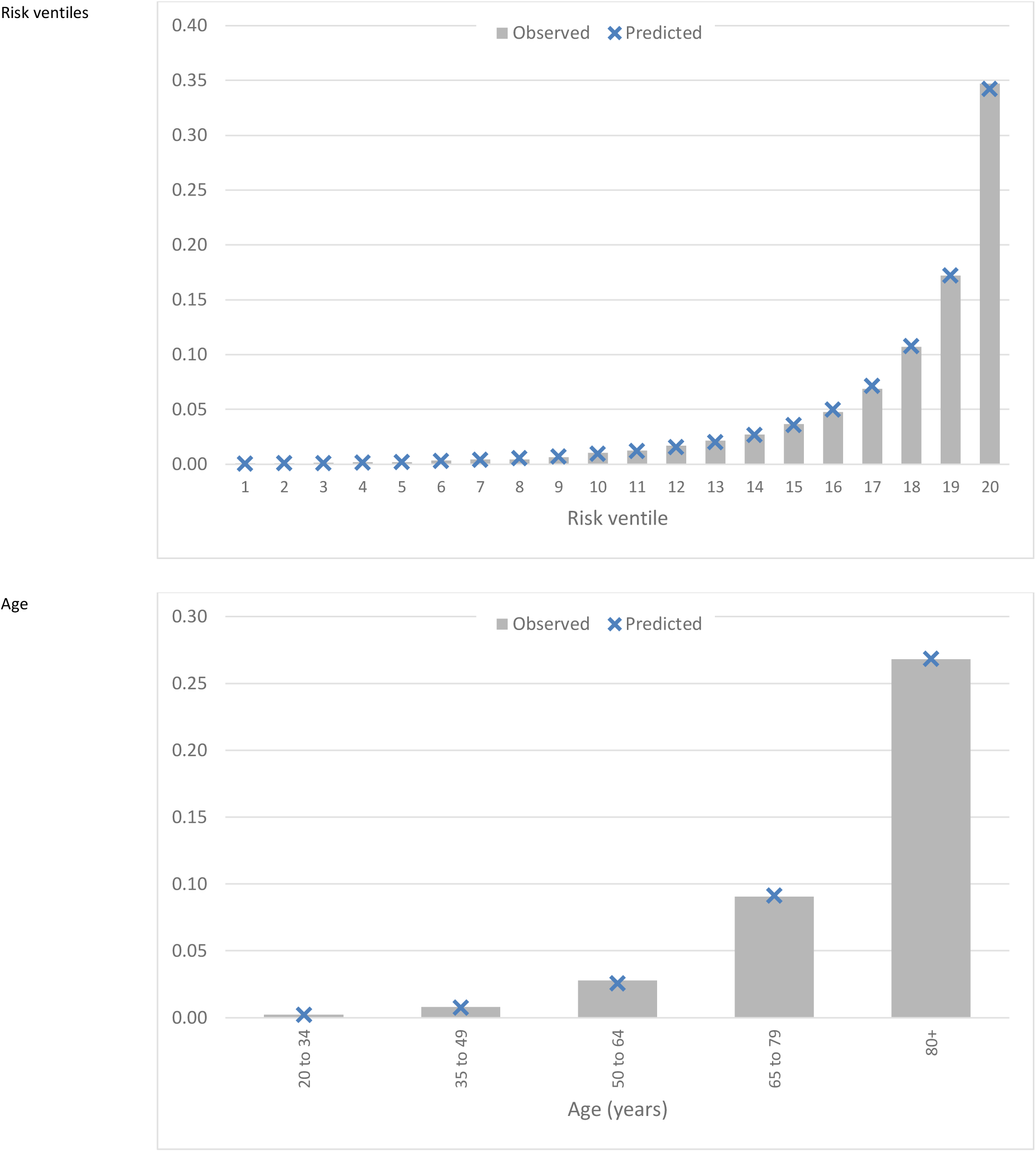
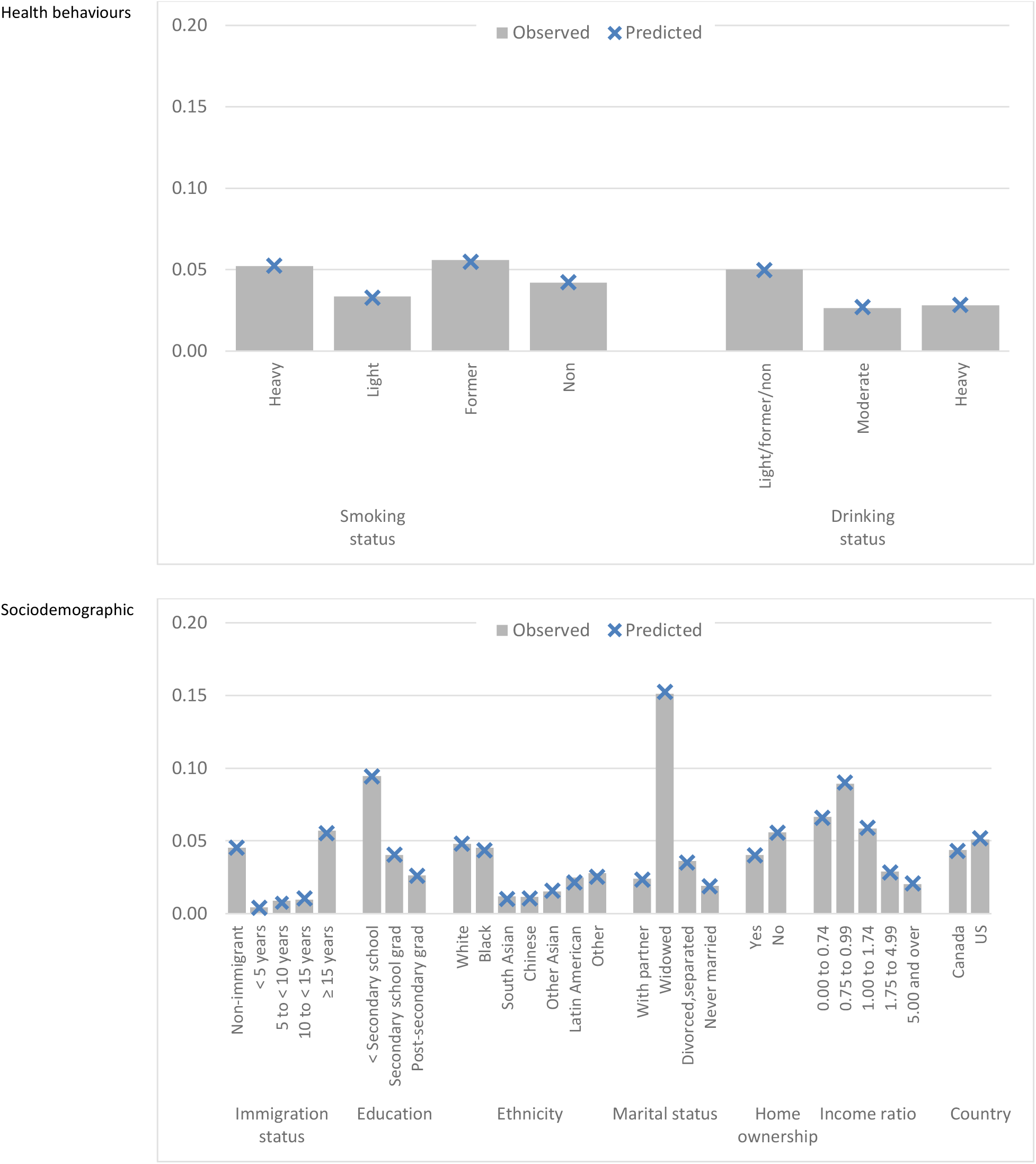
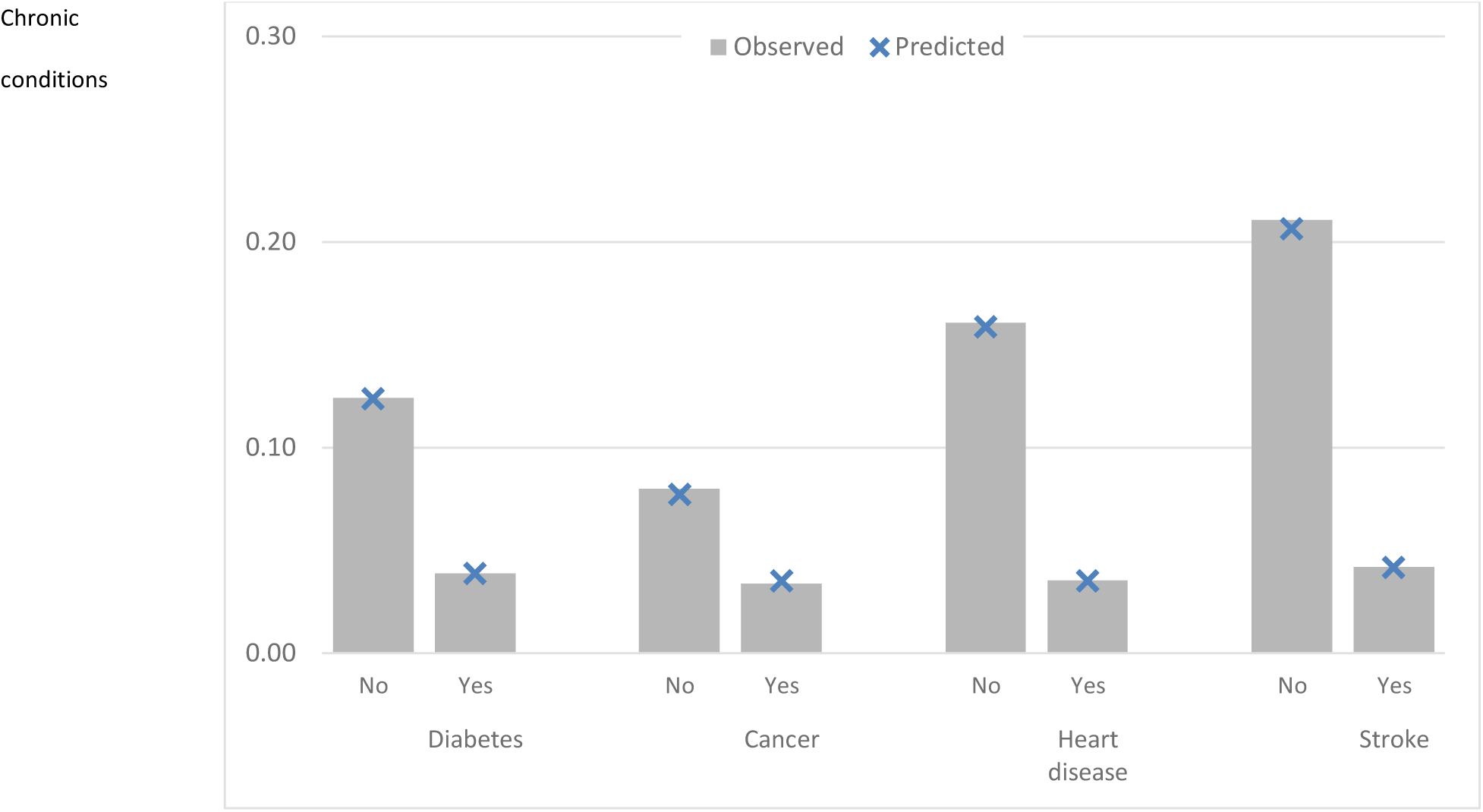

### Additional file 11. Observed versus predicted by subgroups – males external validation, US and Canada combined

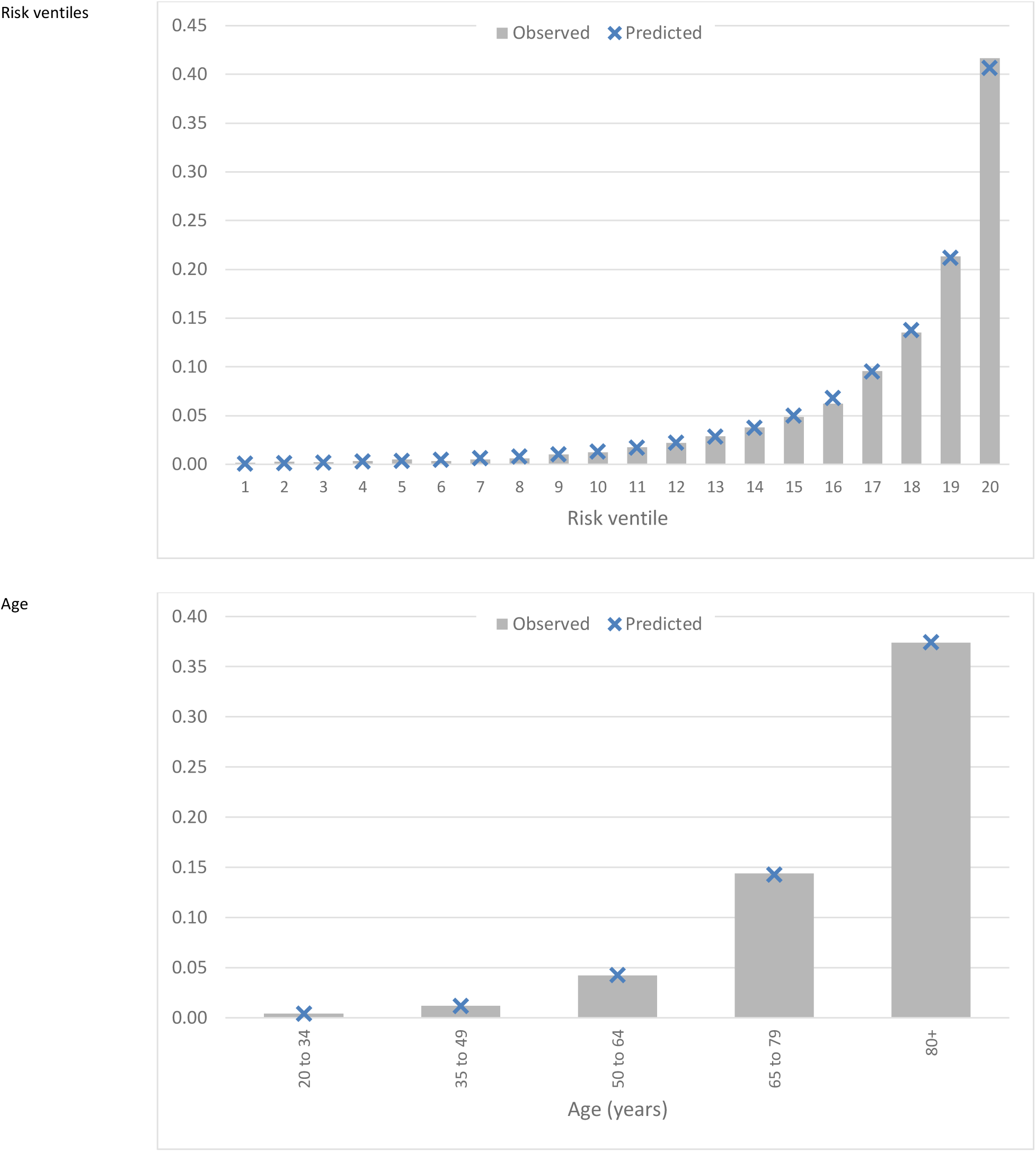
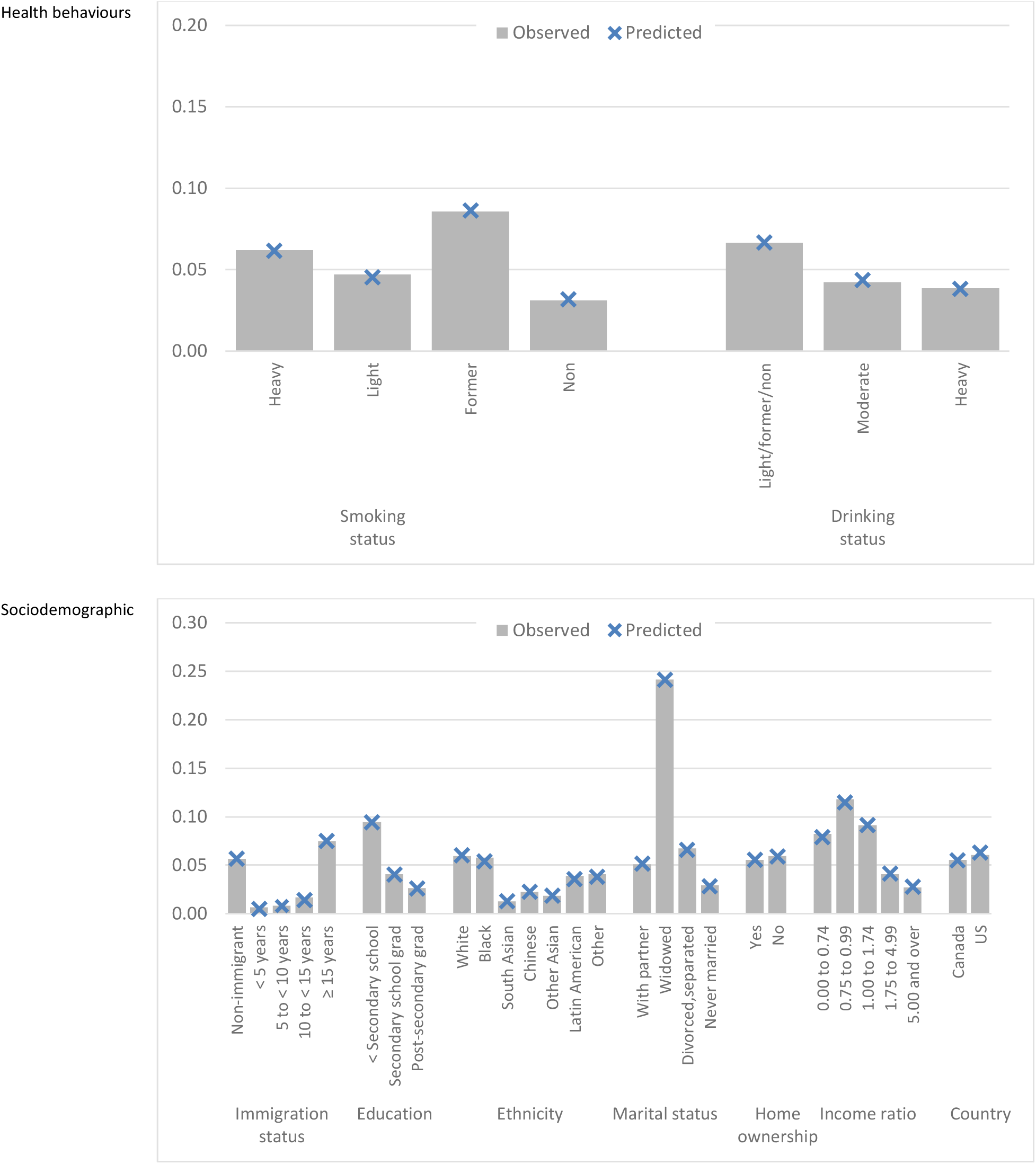
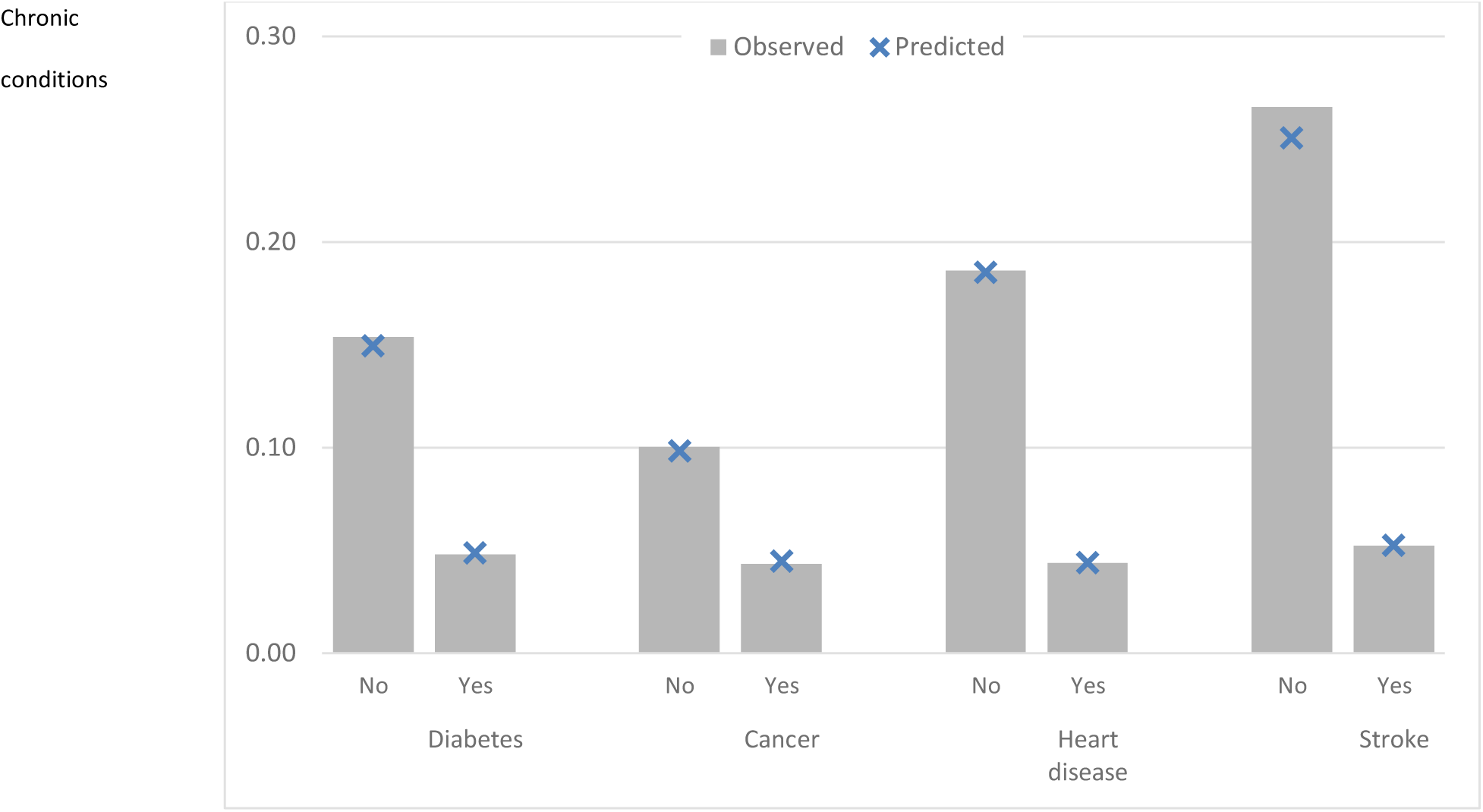

### Additional file 12. GitHub repo README.MD file

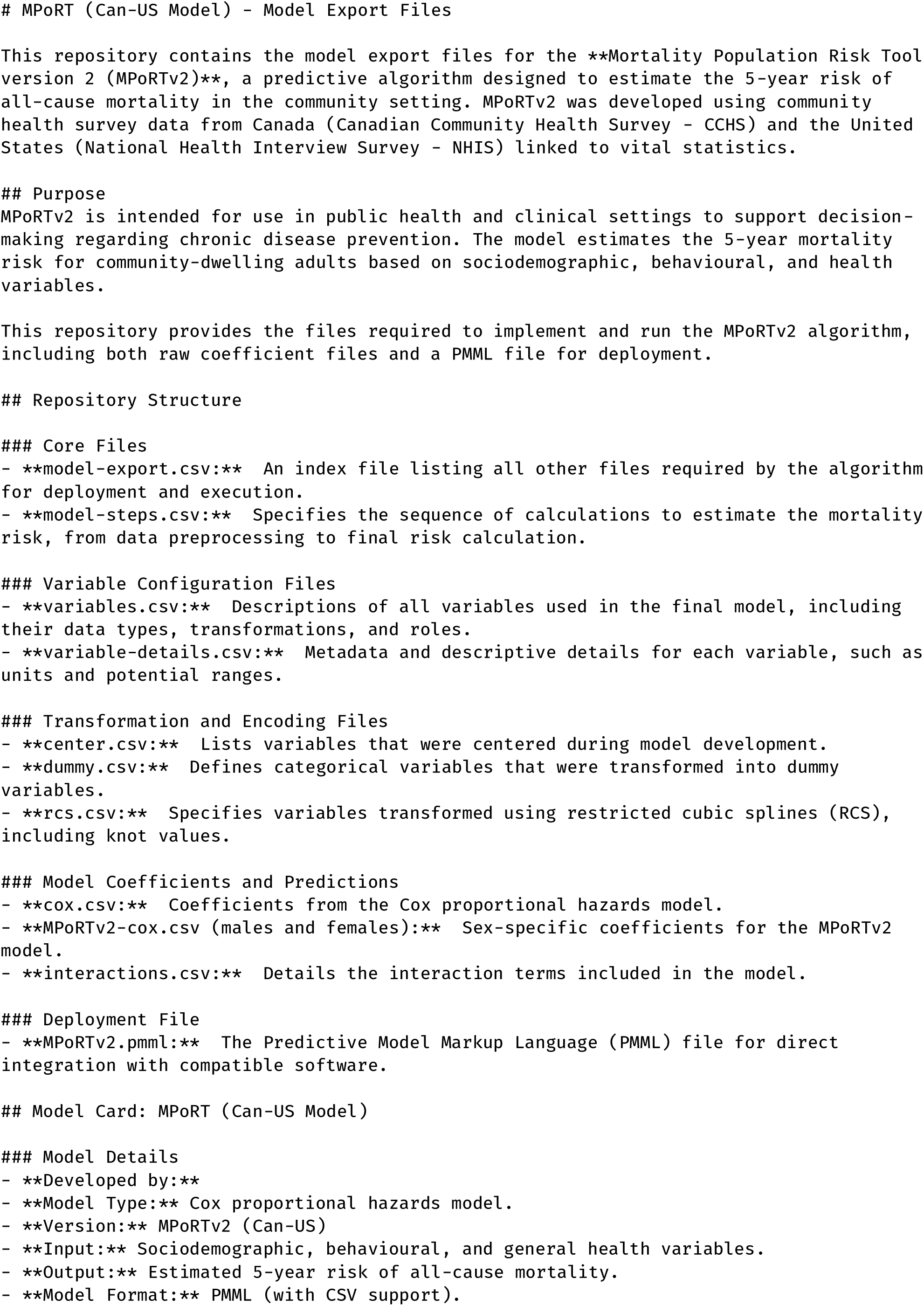
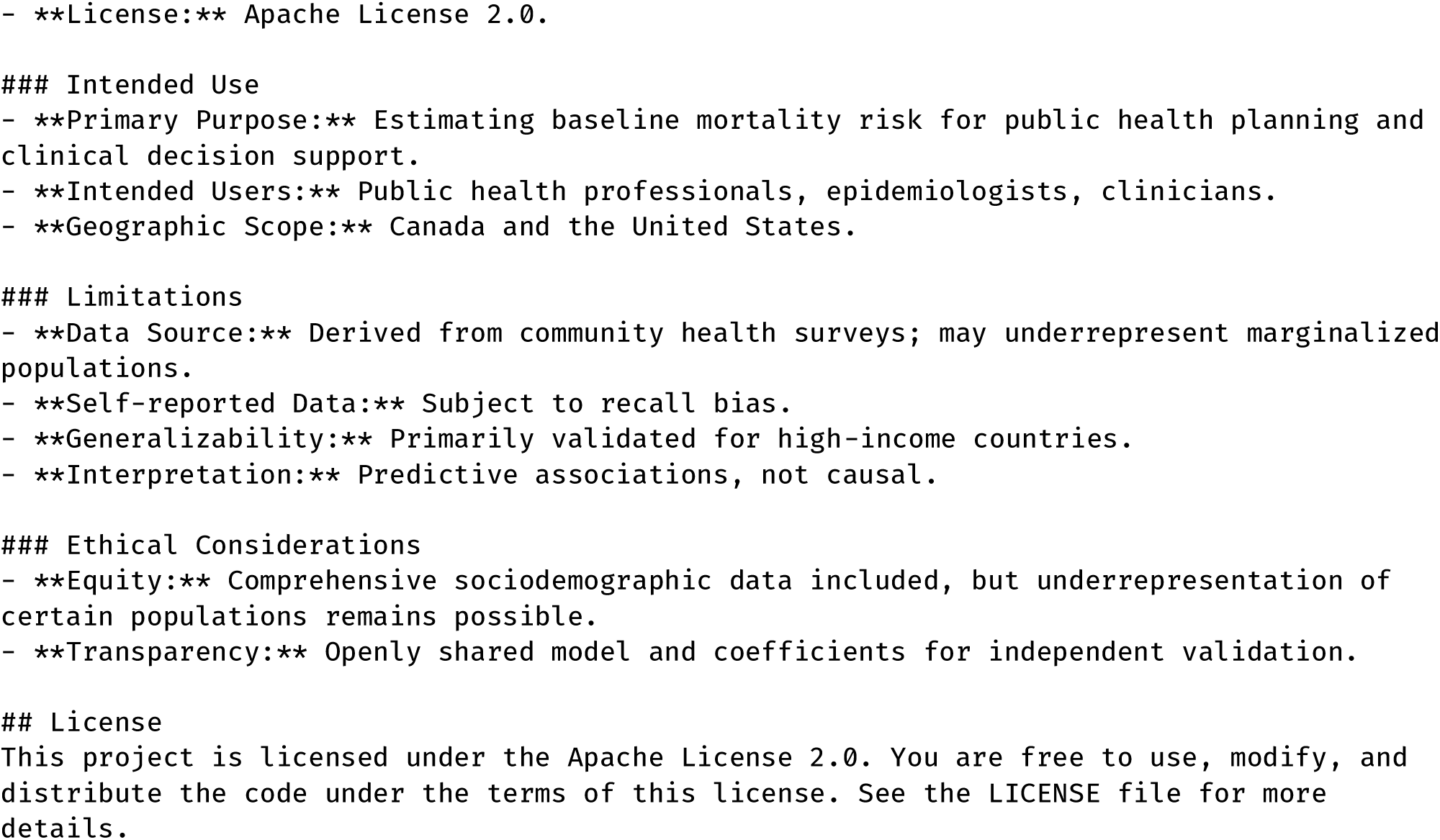

